# Persistent low-level variants in a subset of HCMV genes are highly predictive of poor outcome in immunocompromised patients with cytomegalovirus infection

**DOI:** 10.1101/2022.09.12.22279849

**Authors:** Cristina Venturini, Julia M Colston, Oscar Charles, Timothy Best, Claire Atkinson, Calum Forrest, Charlotte Williams, Kanchan Rao, Austen Worth, Doug Thorburn, Mark Harber, Paul Griffiths, Judith Breuer

**Affiliations:** Infection, Immunity&Inflammation, Institute of Child Health, University College London, London, UK; North Bristol NHS Trust, University Hospitals Bristol and Weston NHS Foundation Trust, Bristol, UK; Great Ormond Street Hospital for Children NHS foundation trust, London, UK; Division of Infection and Immunity, Institute for Immunity and Transplantation, University College London, London, UK; UCL Genomics, University College London, London, UK; Department of Bone Marrow Transplant, Great Ormond Street Hospital for Children NHS foundation trust, London, UK; Department of Immunology, Great Ormond Street Hospital for Children NHS foundation trust, London, UK; Royal Free London, NHS Foundation Trust, London, UK

## Abstract

Human cytomegalovirus (HCMV) is the most common and most serious opportunistic infection after solid organ (SOT) and haematopoietic stem cell transplantation (HSCT). There is considerable interest in using virus sequence data to investigate and monitor viral factors associated with the clinical outcome, including failure to respond to available antiviral therapies.

To assess this, we used target-enrichment to deep-sequence 16 paediatric patients with HSCT, SOT or primary immunodeficiency of whom 9 died with HCMV and 35 infected SOT adult recipients of whom one died with HCMV.

We first showed that samples from both groups have fixed drug-resistance mutations and mixed infections. Deep sequencing also revealed non-fixed resistance mutations in most of the patients who died (6/9). A machine learning approach identified 10 genes with high within-host variability in these patients. These genes formed a viral signature which discriminated patients with HCMV who died from those that survived with high accuracy (AUC=0.96). Lymphocyte numbers for a subset of 17 patients showed no recovery post-transplant of counts in the five who died.

We hypothesise that the viral signature identified in this study may be a useful biomarker for poor response of HCMV to antiviral drug treatment and indirectly for poor T cell function, potentially identifying early, those patients requiring non-pharmacological interventions.

## INTRODUCTION

Human cytomegalovirus (HCMV; human herpesvirus 5) is a member of the *Betaherpesvirinae* subfamily with a worldwide seroprevalence of between 18-100% (1,2). Higher prevalence is linked to lower socio-economic status and older age. HCMV is usually a benign viral infection in immunocompetent individuals, however, it has been shown to be a significant cause of morbidity and mortality in immunosuppressed patients (3,4).

Given that HCMV is the most common and most serious opportunistic infection in these patients, strategies for prevention as well as treatment are of paramount important for transplant clinical success and outcome. Several therapies exist for prophylaxis, pre-emptive therapy and/or treatment of HCMV (5). Treatment with ganciclovir (GCV), foscarnet (FOS), cidofovir (CDV) or letermovir has improved outcomes (6–8), although late resistance often occurs (9). Despite excellent outcomes for most haematopoietic stem cell transplants (SCT) and solid organ (SOT) transplant recipients with HCMV, severe life threatening disease can develop in approximately 20% (7) to 50% of cases (10). Increased use of next generation sequencing (NGS) has associated the presence of fixed drug mutations and mixed infections with poorer outcomes (11–14). To further investigate the pathogenesis of life-threatening HCMV in transplant recipients we analysed 51 patients with refractory HCMV viraemia defined as persisting with less than 0.5 log reduction for three weeks or more despite antiviral treatment (15). Sixteen were HCMV-infected paediatric patients with HCST, SOT or primary immunodeficiency, of whom 9 died with HCMV disease. The other 35 were HCMV-infected SOT adult recipients, of whom one died with HCMV disease. Using a machine learning approach, we identified an HCMV molecular signature which discriminates between patients with different clinical outcome.

## RESULTS

### Patients’ characteristics

We analysed two different cohorts: 16 retrospectively identified paediatric patients from Great Ormond Street hospital for Children (GOSH) with primary immunodeficiency syndromes (PIDs) or HSCTs (the characteristics of patients are shown in Table 1) and 35 adult SOT recipients from Royal Free Hospital London. All had HCMV viraemia persisting with ≤0.5 log reduction despite antiviral treatment for 21 days or longer, which has been defined as refractory (15).

No patients received prophylaxis against HCMV, although all SCTs received standard acyclovir prophylaxis against alpha-herpesviruses. Pre-emptive antiviral treatment for HCMV was initiated at first detection in the PIDs, when viraemia exceeded 1000 IU/ml in the SCT recipients and 3000 IU/ml in the SOTs. First line therapy was ganciclovir in the SOTs and PIDs and foscarnet in the SCT recipients.

We stratified patients into two groups: a poor outcome group, defined as those who died with HCMV viraemia (n=9) and a good outcome group defined as patients who cleared their HCMV (n=42).

### Deep sequencing metrics

A total of 149 samples from 51 patients (1-9 samples per patient) were mapped to the HCMV reference strain Merlin genome (NC.006273). Samples were included in further analysis if they reached >95% coverage of the reference strain and a mean read depth (MRD) of >10x. MRDs ranged from 10x to 1407x (mean 143x). Details of mapping statistics and quality are shown in Supplementary Table 1.

### Drug resistance mutations

Resistance to antiviral drugs has been associated with poor outcome. To examine this in our patient cohort we annotated sequence data using a comprehensive database, derived from published literature (16), to identify fixed resistance in the UL97 (serine/threonine protein kinase) and the UL54 (DNA polymerase) genes, which are the targets of the anti-HCMV drugs GCV (UL97 and UL54) and FOS (UL54) used here. Fixed resistance (>50% frequency) mutations were identified in 5/42 (11.9%) in the good outcome group and 3/9 (33.5%) in the poor outcome group (*X*^2^=2.57, p-value=.19).

### Poor outcome is not associated with multiple HCMV strain infection

Mixed HCMV infections have been previously linked with poor clinical outcome (12). To investigate this in our two cohorts, we first calculated genome-wide within-host diversity (π) for each sample (12) (Figure 1A). HCMV sample diversity separated into higher and lower diversity groups, with the majority having low within-host diversity. The distribution of diversity showed bi-modality (p-value=0.016, first peak/mode estimated at 0.0015 and second peak estimated at 0.0077, Figure 1 – Supplementary figure 1A). The two estimated peaks were used to fit two Gaussian distributions to the data (Figure 1 – Supplementary figure 1B) which crossed at >0.005. Based on previous analyses (12) we used this value as the cut-off above which infections were considered as potentially mixed. We reconstructed haplotypes in all 18 patients with at least one sample with diversity greater than 0.005. 14 patients were confirmed to have multiple strains where haplotypes differed by at least 2kbp with the minor haplotypes present at >5% frequency (17) (Figure 1B). Mixed infections were not predictive of clinical outcome (26% of patients with good outcome presented with multiple strains versus 33% with poor outcome, *X*^2^=0.19, p-value=.66).

**Figure 1A:**
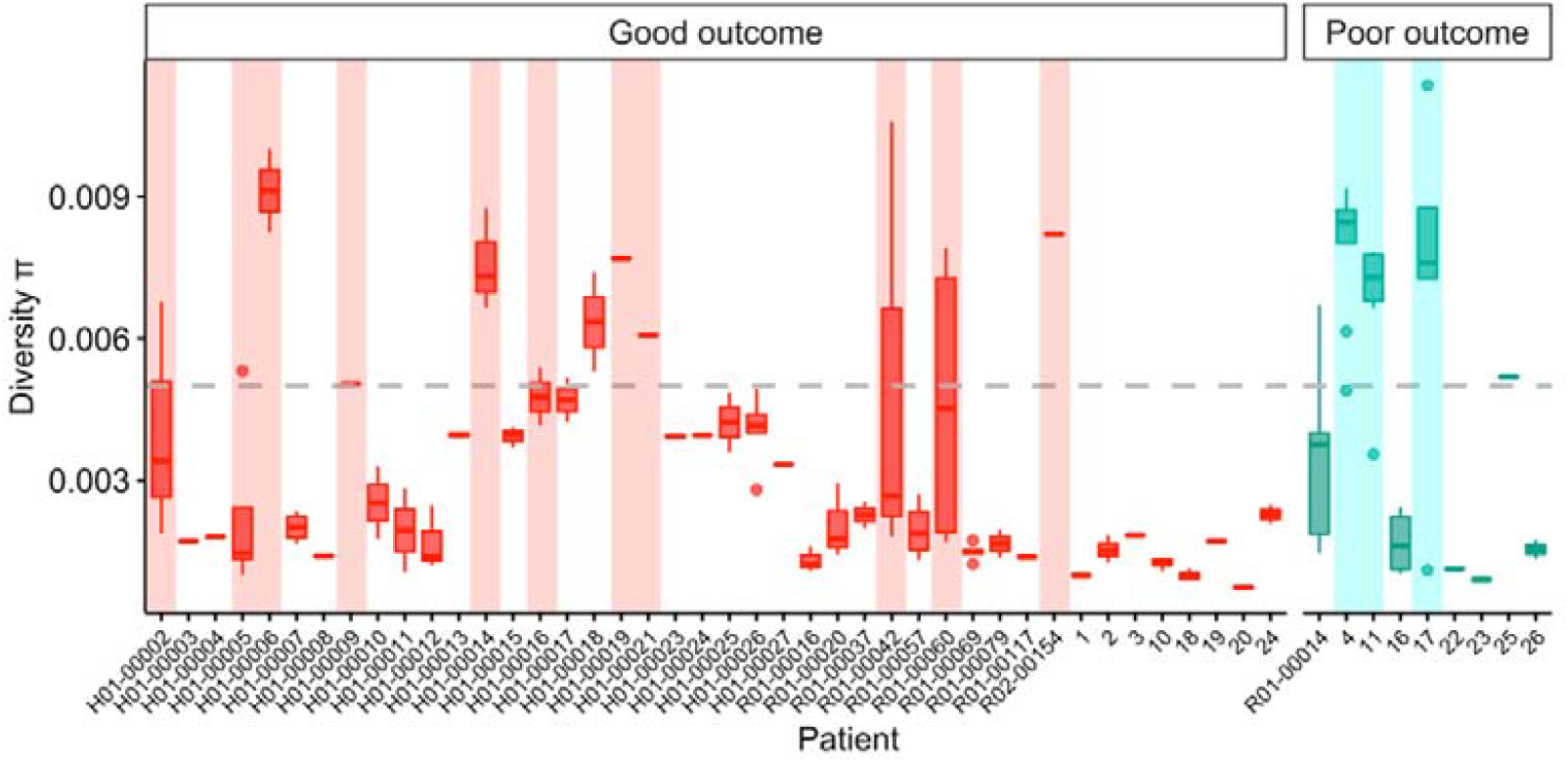
Diversity values for all patients. Each patient (x-axis) had longitudinal samples (box plots) showing diversity values (y-axis). Patients with good clinical outcomes are represented in red and those with poor outcomes are in turquoise. Dash grey line represents the chosen cut-off of 0.005 for investigating mixed infections. Patients with transparent rectangles were confirmed mixed infections.

**Figure 1B:**
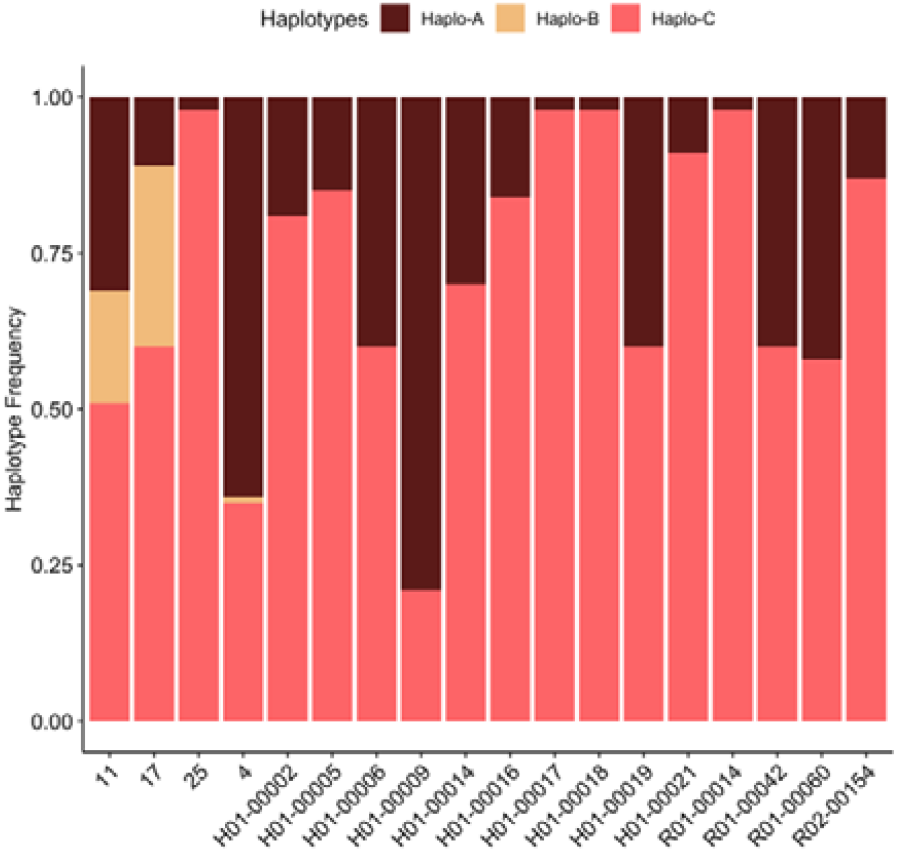
Reconstructed haplotype abundances for 18 patients suspected from their diversity scores of having mixed infections. For each patient haplotypes were reconstructed; here, for representative purposes, we only show one sample for each patient. Minor haplotypes occurring at <5% were discounted (17).

### Low-level variation in all HCMV genes and genes selection

Deep sequencing also revealed low frequency (<50%) GCV and FOS resistance mutations in 6/9 of the poor outcome patients, two of whom also had mixed infections and 2/42 in the good prognosis group, none of whom had mixed infections (Table 2, *X*^2^ =21.47, p-value=.00001). Most variants occurred at frequencies <15% (median frequency 13.45) (Table 2 – figure supplement 1). In all patients in the poor outcome group with multiple longitudinal samples, low frequency resistance mutations persisted in multiple samples, with the majority failing to rise to fixation. In contrast, low level resistance variants present in the two patients in the good outcome group were at low frequency at the first-time point rising to fixation in later samples (Table 2 – figure supplement 1).

**Table 2:**
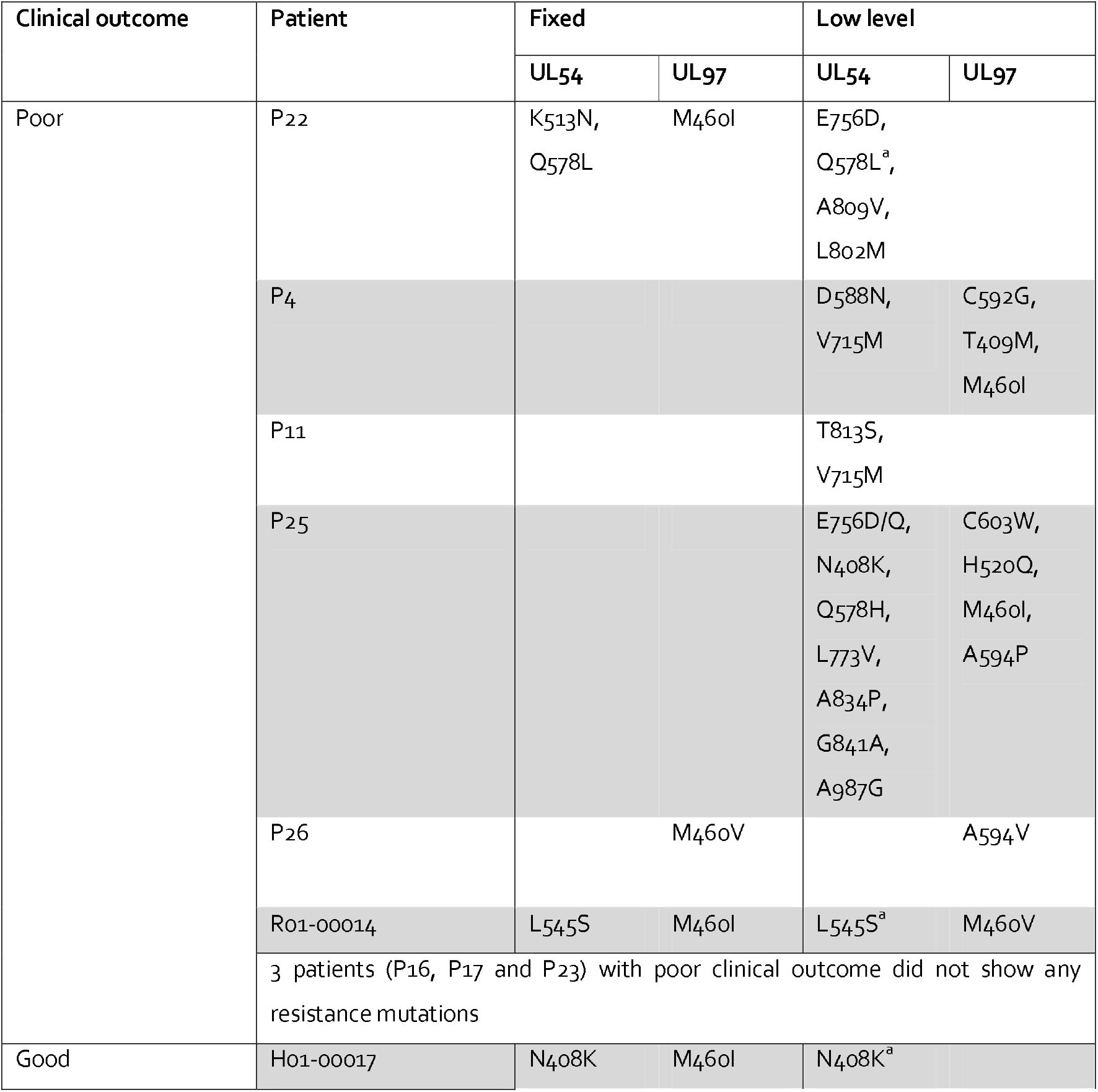
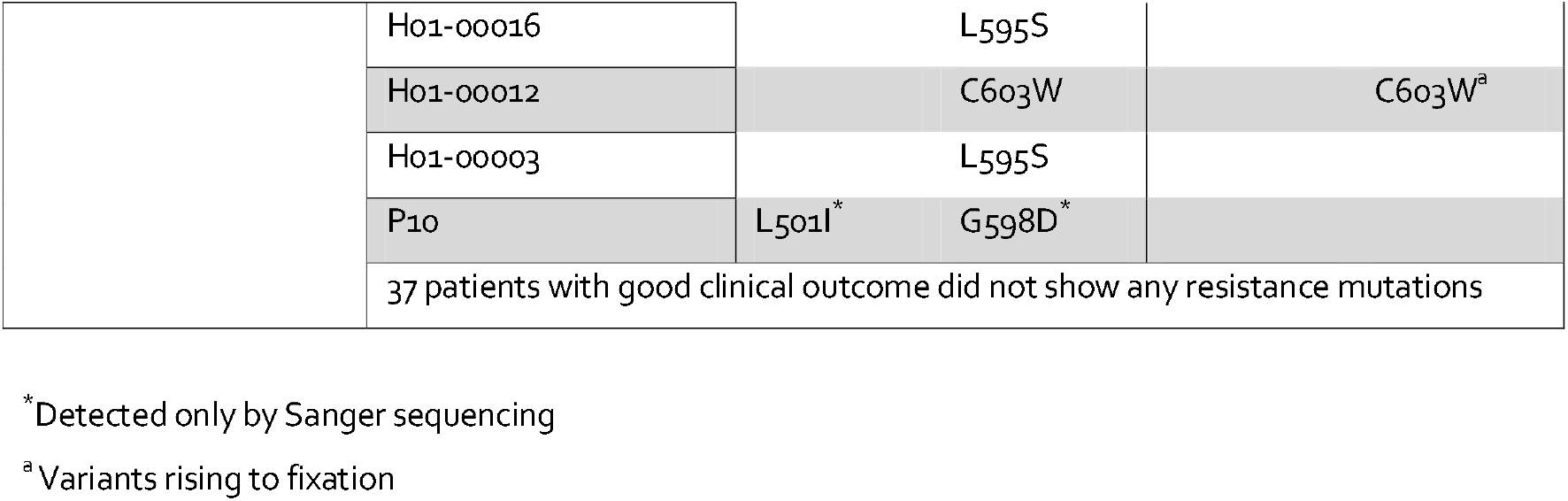
All detected drug resistance mutations (FC >=2)

To determine whether low level HCMV mutations (or minority variants, MVs) were confined to positions coding for antiviral resistance, we investigated all non-synonymous (NS) MVs across the genomes in the single infections (initially excluding all patients with mixed infection to simplify mapping). In the whole population, there was no evidence of clustering (Figure 2) by open reading frame, with NS MVs located apparently randomly across the HCMV genome.

**Figure 2:**
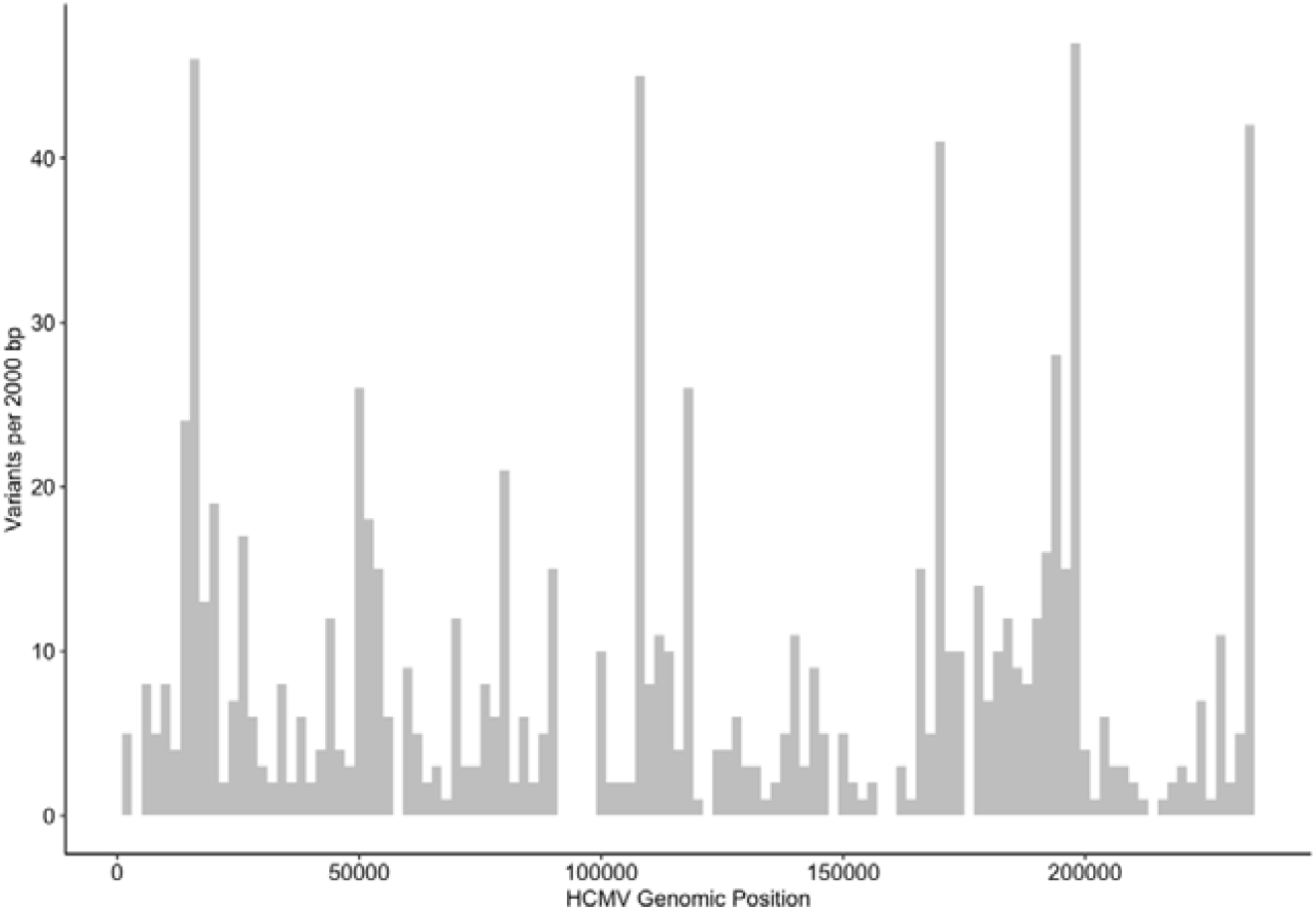
Minority variants distribution across the HCMV genome in all samples

We then compared samples from patients with single infections with different clinical outcomes. We ranked the HCMV genes that discriminated patients who died from patients who survived using machine learning methods. To do this, we implemented a gene selection algorithm (using chi-squared statistics) to evaluate the importance of the presence of MVs in a specific gene. The gene selection process identified 10 genes with high K score (K score > 8) and significant p-values (p-values < 0.005 and adjusted p-value <0.5) (Figure 3).

**Figure 3:**
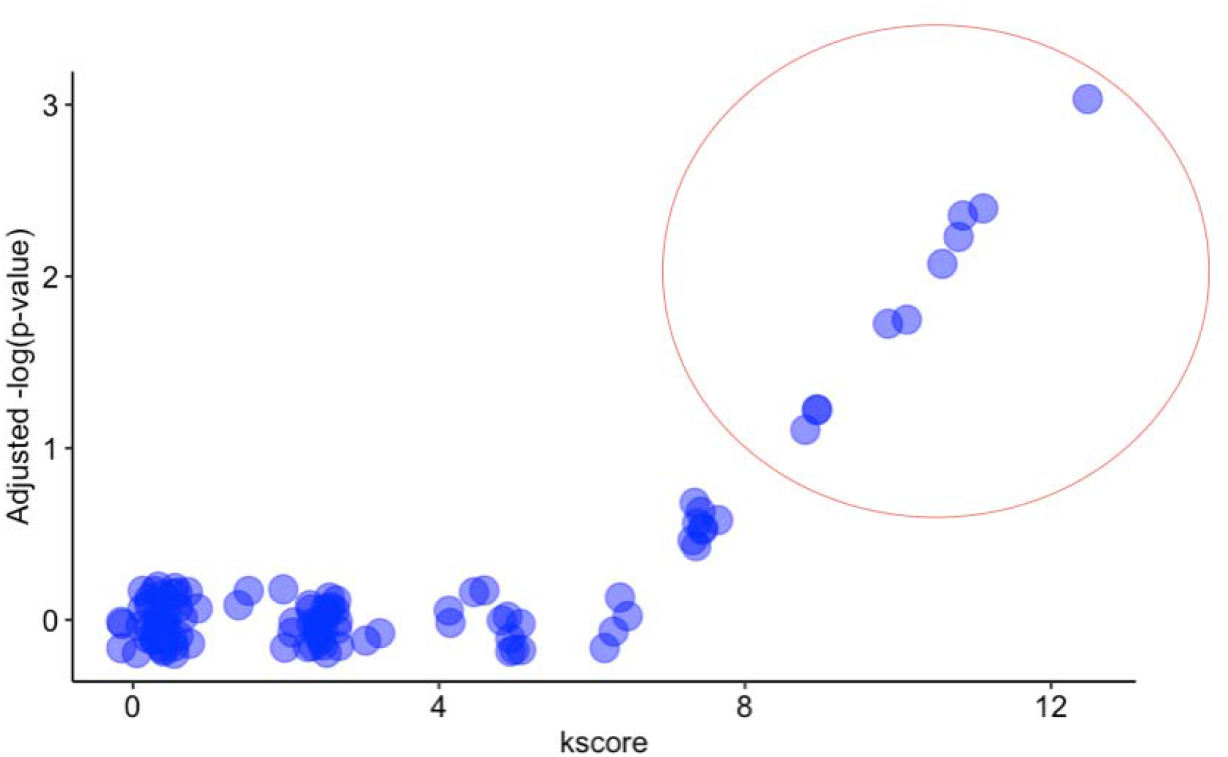
Feature selection results: x-axis shows the k-score and y-axis the -log(p-value) adjusted for multiple testing. We selected the top 10 genes with k-score >8 and p-values <0.005.

All ten genes showed more within-host variability in the poor outcome groups compared to the good outcome group. No gene showed the opposite trend of being more variable in the good outcome group compared to samples from patients who died.

The variable genes in the patients with poor outcome included the polymerase gene (UL54) and the serine/threonine protein kinase (UL97), which are both already known for their association with drug resistance. In addition, we identified genes coding for glycoproteins (envelope gp such as UL74, gO and UL75, gH; immediate early gp, UL37; membrane gp UL7), membrane proteins UL121 and UL8 and the genes coding for the uncharacterized proteins UL20 and UL11, the latter of which plays a role altering host immune response by modulating T-cell function.

We focussed on NS MVs as these gave better discrimination between poor and good outcome groups than non-synonymous and synonymous mutations combined, for all genes, bar UL11, UL7 and UL97 (Table 3).

**Table 3:** Genes are ranked by k-score and p-value. Column “All” shows k-score and p-values for analysis including both synonymous and non-synonymous variants.

| Ranking | Gene | NS K-score/p-value | All K-score/p-value |
| --- | --- | --- | --- |
| 1 | UL54 | 12.5/4.07e-04 | 3/3.89e-03 |
| 2 | UL20 | 11.2/8.18e-04 | 0.6/4.1e-01 |
| 3 | UL11 | 10.6/1.08e-03 | 11/8.58e-04 |
| 4 | UL8 | 10.6/1.08e-03 | 2/0.15 |
| 5 | UL37 | 10.5/1.18e-03 | 0/1 |
| 6 | UL121 | 10/1.56e-03 | 8/3.89e-03 |
| 7 | UL75 | 10/1.56e-03 | 1.8/1.77e-01 |
| 8 | UL7 | 8.8/2.98e-03 | 18/1.4e-05 |
| 9 | UL97 | 8.8/2.98e-03 | 13/2.57e-04 |
| 10 | UL74 | 8.7/3.09e-03 | 2/1.57e-01 |

### Viral signature in HCMV samples from patients with poor clinical outcome

To assess the predictive power of using the ten gene viral signature, we calculated the True Positive Rate (TPR, sensitivity) and the False Positive Rate (FPR, 1-Specificity). We then built the Receiver Operating Characteristics (ROC) curve and calculated the area under the curve (AUC) which provides an aggregate measure of the performance of the model. We compared two models: a) a full model including presence of NS MVs in the 10 signature’s genes; and b) a drug resistance gene model, where we only included NS MVs in UL54 and UL97. The full model had an AUC of 0.96 and was statistically more discriminatory than the model with only resistance genes (p-value < 0.001, Anova, Figure 4A). We used the full model to calculate, and plot estimated probabilities of having a poor clinical outcome (Figure 4B).

**Figure 4A:**
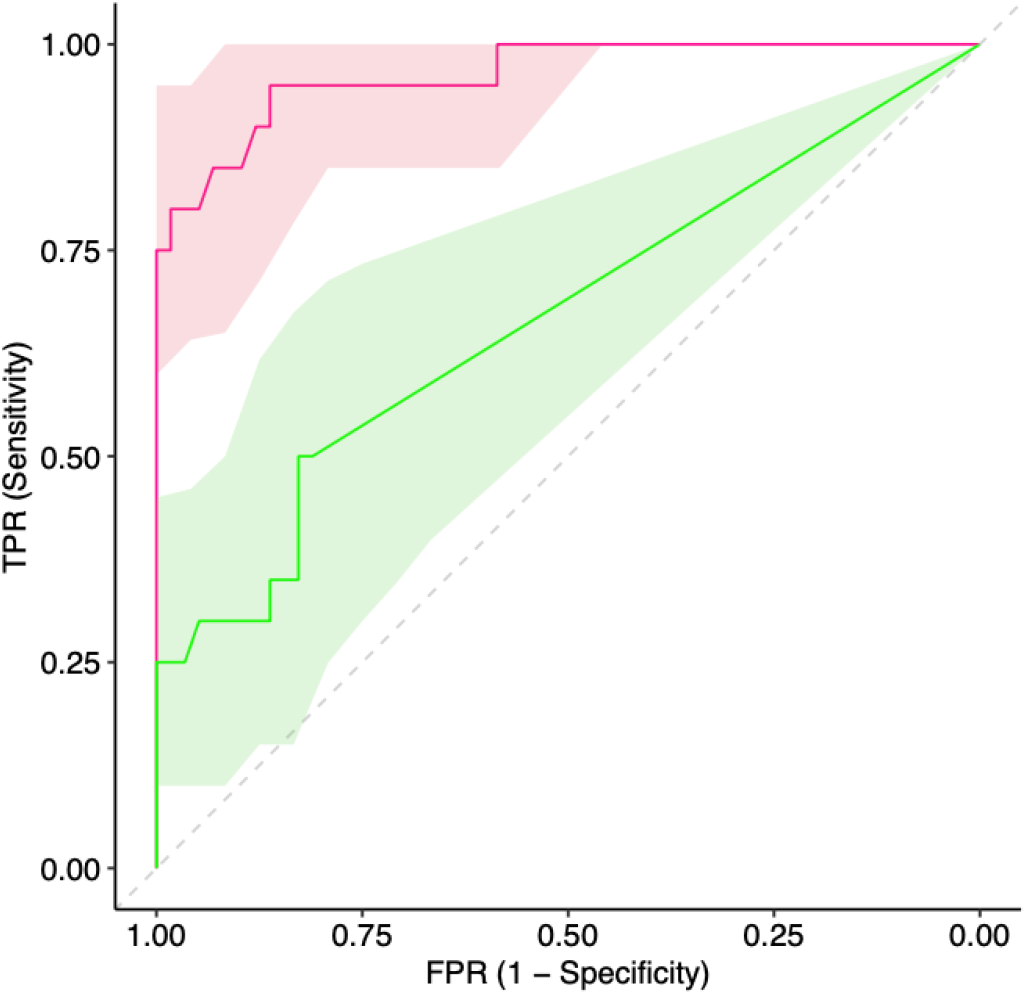
ROC curves with confidence intervals (95%) for two predictive models discriminating between samples from patients who died and survivors. AUC for the full model (including MVs in the 10 candidate genes) was 0.96 (red ROC curve). AUC for the drug resistance genes model (including genes UL54 and UL97) was 0.65 (green ROC curve).

**Figure 4B:**
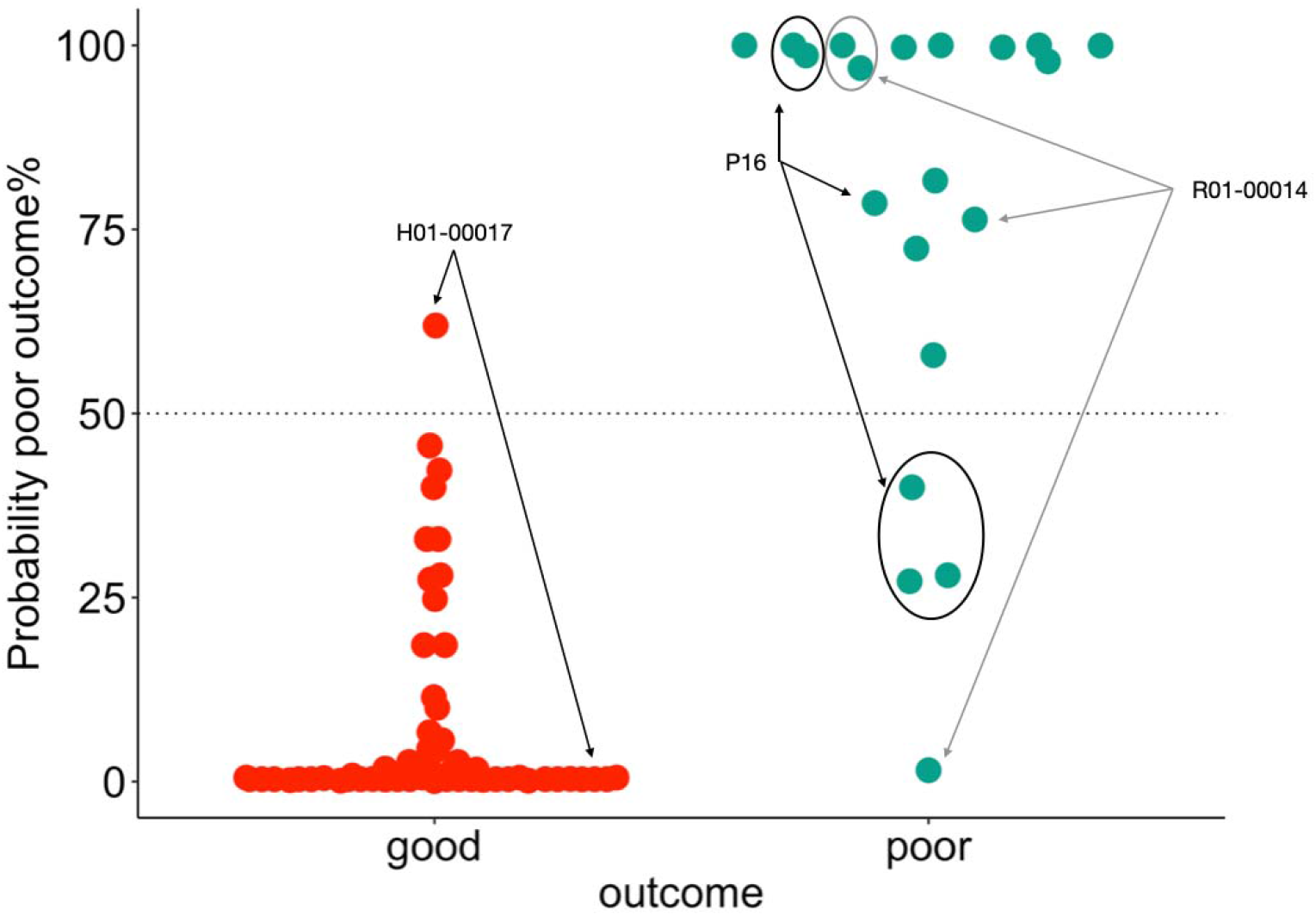
Estimated probabilities for each sample in the two groups (red for survivors, turquoise for patients who died) to be classified as a patient from the poor outcome group. Arrows and circle indicate patients with at least a sample which was misclassified by the model.

Only 5 samples from 3 patients were misclassified by the full model (Table 4, Figure 4B). One sample from the good outcome group was classified as “poor outcome”. The patient (H01-00017) was an adult who received a kidney transplant with two episodes of post-transplantation HCMV viraemia (one lasting 28 days and one 149 days). Both samples were taken during the second episode. The first sample taken at 77 days after transplant and 20 days from the start of the second episode of HCMV viraemia and associated antiviral treatment, had a probability of 62% of poor outcome. However, his second sample taken 25 days later, i.e. after 45 days of continuous treatment, had a probability of poor outcome of 0%, despite the rise to fixation of a low level GCV resistance mutation. This patient was in fact one of only two in the good outcome group with multiple resistance mutations, one fixed and another rising to fixation in the second sample.

**Table 4:**
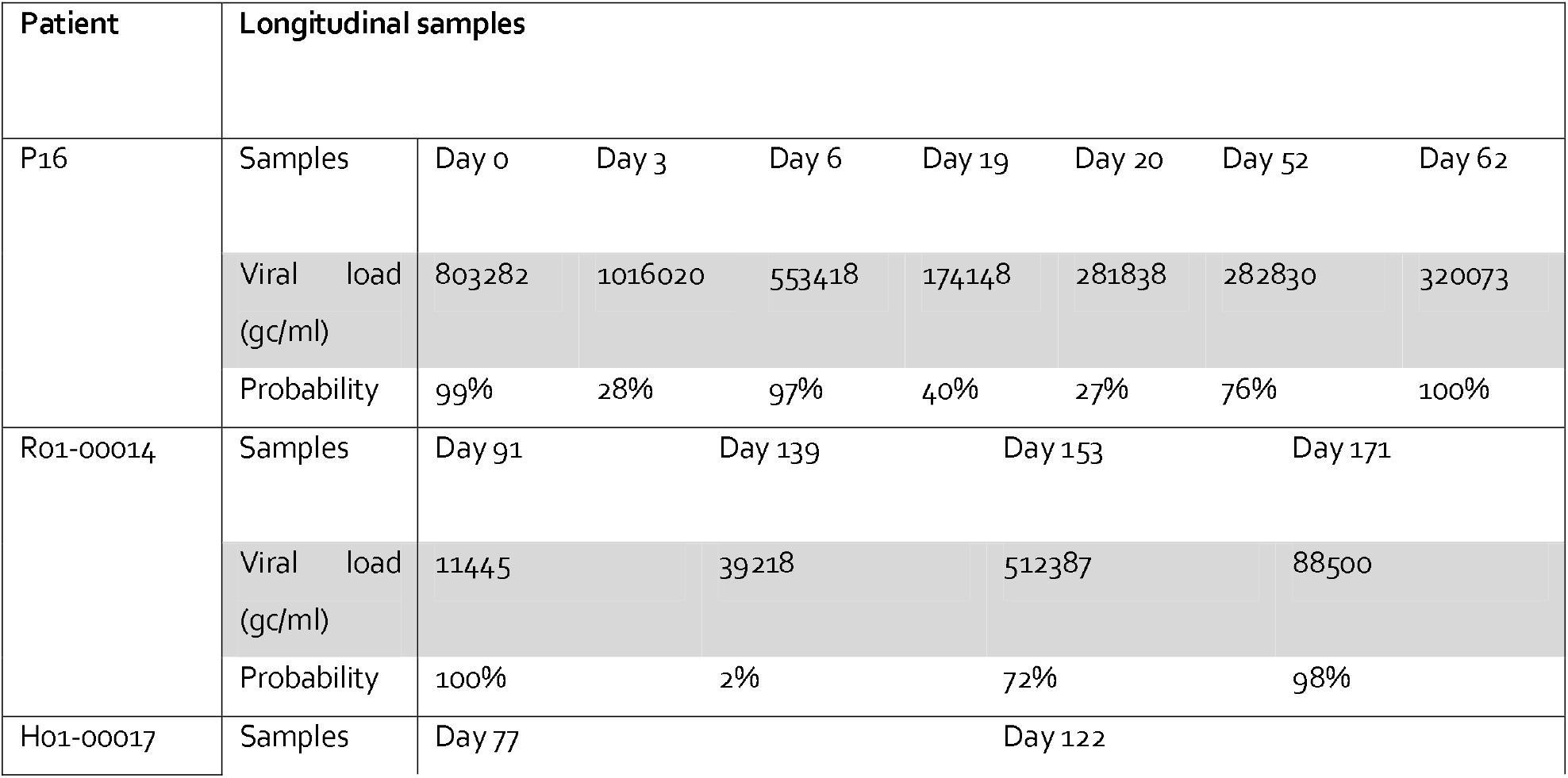
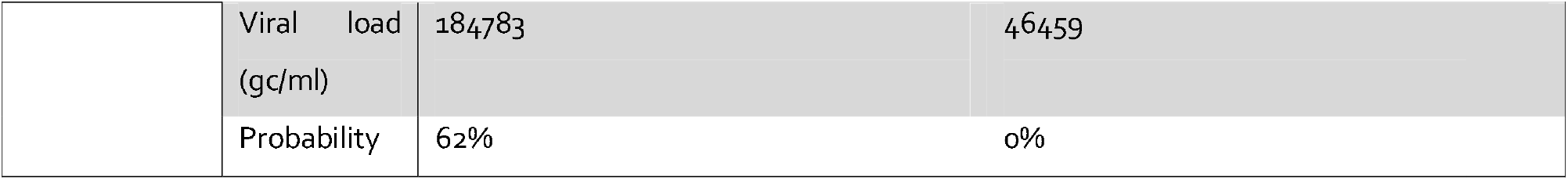
Patients with samples classified as “ambiguous”.

| Patient | Longitudinal samples |  |  |  |  |  |  |  |
| --- | --- | --- | --- | --- | --- | --- | --- | --- |
| P16 | Samples | Day 0 | Day 3 | Day 6 | Day 19 | Day 20 | Day 52 | Day 62 |
|  | Viral load (gc/ml) | 803282 | 1016020 | 553418 | 174148 | 281838 | 282830 | 320073 |
|  | Probability | 99% | 28% | 97% | 40% | 27% | 76% | 100% |
| Ro1-00014 | Samples | Day 91 | Day 139 | Day 153 | Day 171 |  |  |  |
|  | Viral load (gc/ml) | 11445 | 39218 | 512387 | 88500 |  |  |  |
|  | Probability | 100% | 2% | 72% | 98% |  |  |  |
| Ho1-00017 | Samples | Day 77 | Day 122 |  |  |  |  |  |

|  |  |  |  |
| --- | --- | --- | --- |
|  | Viral load<br>(gc/ml) | 184783 | 46459 |
|  | Probability | 62% | 0% |

In the poor outcome group, patient P16 with RAG2 SCID, who died 45 days after starting antivirals for HCMV, had three of seven samples with a probability of <50% (28%, 40% and 27%) of being in the poor outcome group. These three samples were collected at days 3, 19 and 20 with viral loads of respectively 1016020, 174148 and 281838 gc/ml. Treatment with FOS and GCV was started at day 17. The samples with low probability of poor outcome were interspersed with samples taken at days 0 and 6 (viral loads 803282 and 553418 gc/ml respectively) showing probabilities of 90% of being in the poor outcome group. There was no correlation between viral load and probabilities, for example at day of peak viral load (1016020 gc/ml) the probability was 28%. A further two samples taken after day 20 showed probabilities of 76% and 100% of poor outcome. This patient did sadly subsequently die with HCMV infection.

A second patient (R01-00014) who died with HCMV infection also had an apparently low probability (2%) of poor outcome from a sample taken at 139 days post liver transplant. However, another three samples taken at 91, 153 and 171 days post-transplant showed probabilities of 100%, 72% and 98% of poor outcome. Average read depth in all samples were >100x.

We also assessed the predictive power of the signature including single and mixed infections. The full model including mixed infections had a high predictive power (AUC=0.91), albeit lower than the model with only single infections (AUC=0.96), likely due to the difficulty in assembly and calling minority variants where two or more strains are present in a single sample (Figure 4 – supplement figure 1).

### Viral signature over time

To determine how early low-level mutations in the ten sentinel genes can be used to predict a potentially poor outcome, we plotted the probability of being in the poor outcome group for three patients with poor outcome for whom we had multiple longitudinal samples (Figure 5). We also plotted longitudinal data for two patients from the good outcome group who had low-level resistance mutations.

**Figure 5:**
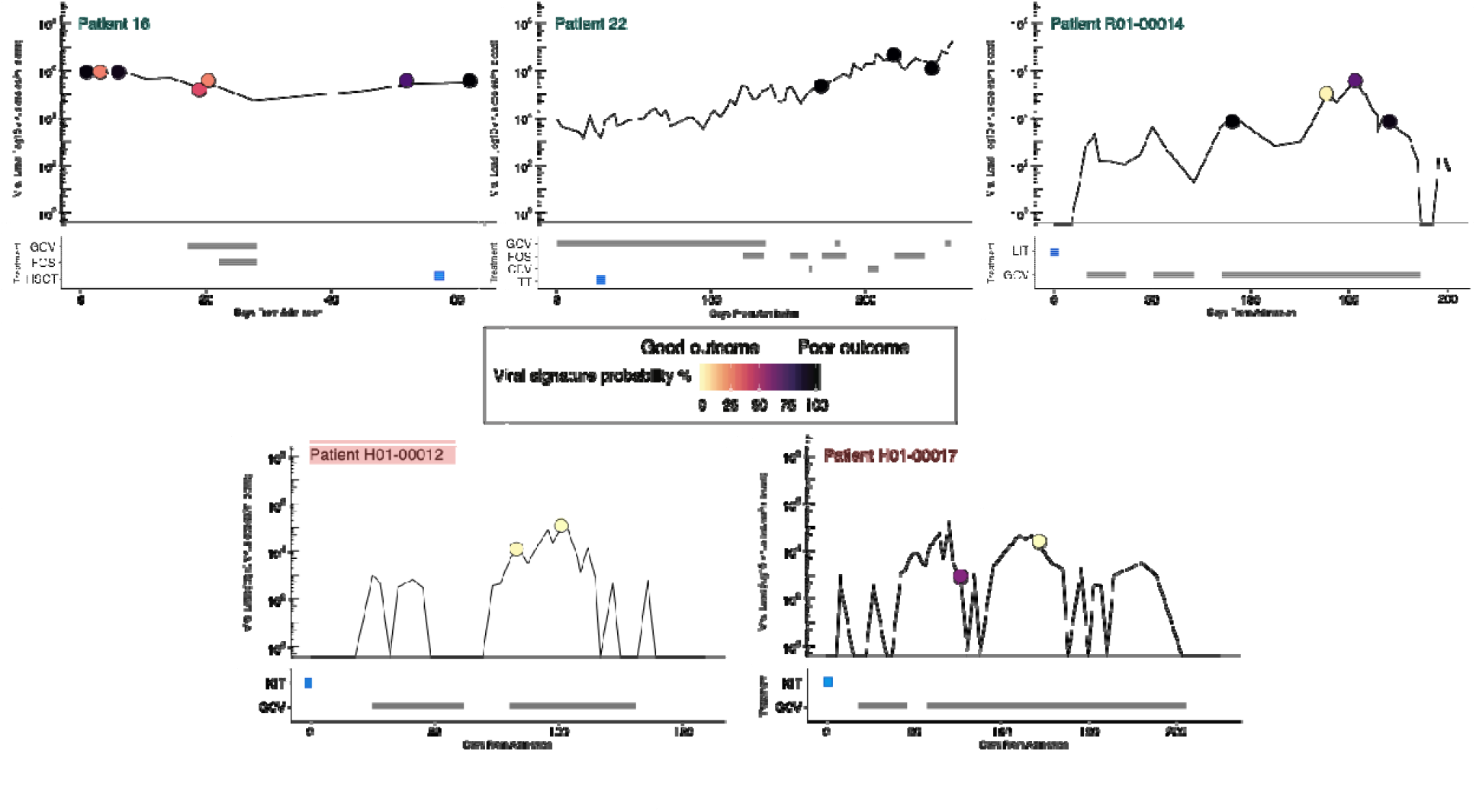
Viraemia, anti-viral therapy and transplant and viral signature probability in patients with longitudinal samples and low-level resistance mutations, including patient 16, 22 and R01-00014 from the poor outcome group and H01-00012 and H01-00017 from the good outcome group. Dots indicate samples sequenced and are coloured based on viral signature probabilities (from yellow, good outcome, to black, poor outcome). Black rectangles indicate anti-viral treatment, and the blue square shows the time of transplant (HSCT for patient 16, liver for patient R01-00014 and thymus for patient 22, kidney for patients H01-00012 and H01-00017).

Since samples from earlier during HCMV infection in patients 22 and R01-00014 with poor outcome were not available for sequencing, it was not possible to determine the earliest time at which the signature appeared. However, samples taken at days 171 (from admission) and 91 (from transplant) respectively i.e. 62 and 109 days before death were positive for the predictive signature. In patient P16 the signature was present as early as 9 days after HSCT.

### Biological significance of the MVs

Most of the HCMV genome is under purifying selection (18), presenting on average a greater proportion of synonymous (S) changes compared to NS and stop codons. Surprisingly five of the ten genes in our viral signature (UL54, UL20, UL121, UL97 and UL74) reversed this trend with greater NS vs S MVs (Table 5). In the genes, NS variants tended to cluster closer together than expected by chance suggesting a functional role. In addition, most of the MVs (63%) mapped to HCMV variable loci identified comparing GenBank sequences. A higher overlap was observed for hypervariable genes (e.g. UL74 (19)) compared with drug resistance genes (e.g. UL54) (Table 5). Clustering of variable residues is a feature of epitopes for which plasticity provides advantages in the face of host immunity. To explore this possibility, we identified known and predicted T cell epitopes from the IEDB database which overlap with amino-acid changes seen in samples from patients who died. We found epitopes in 8/10 genes, which included the 5 genes with greater NS vs S MVs.

**Table 5:**
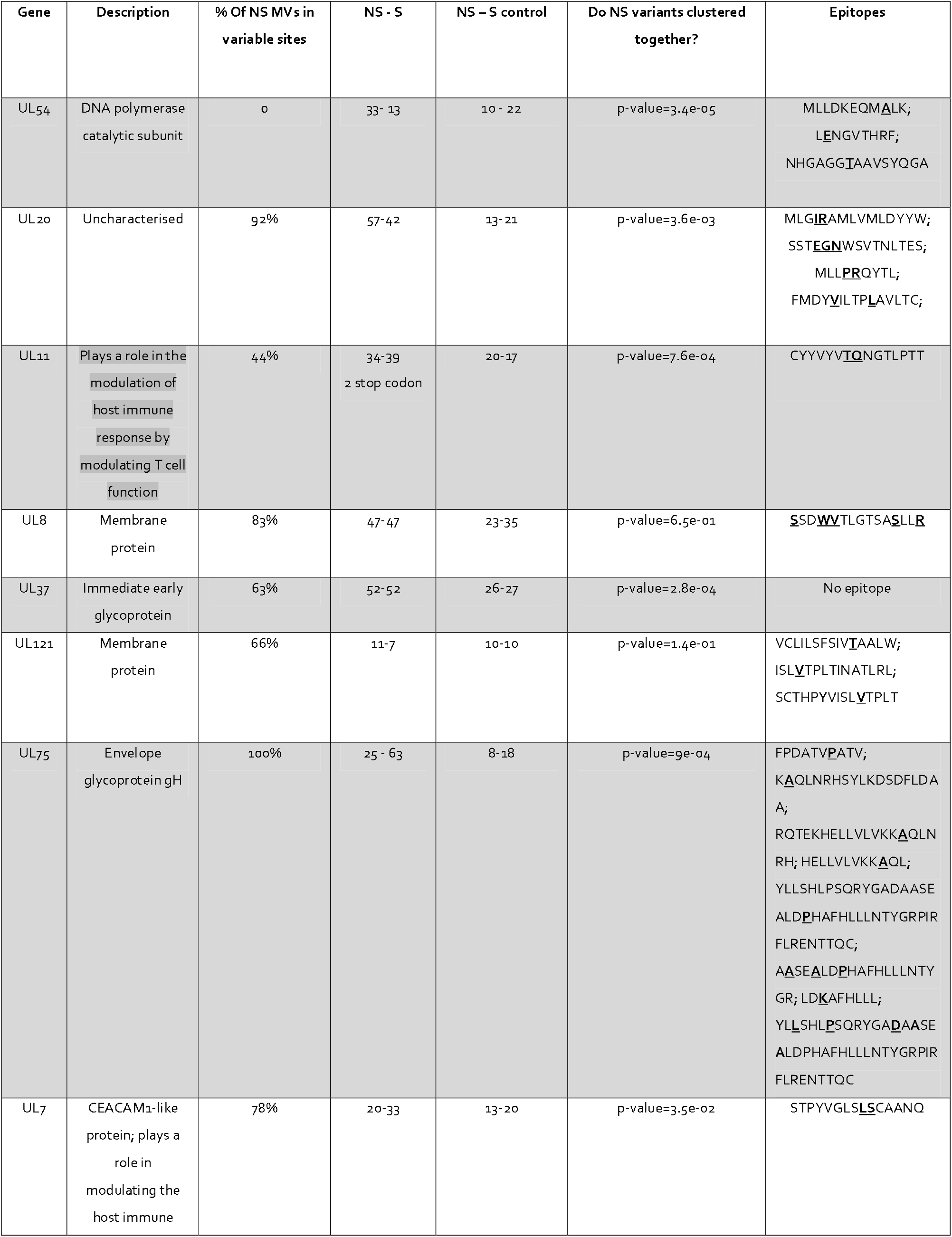
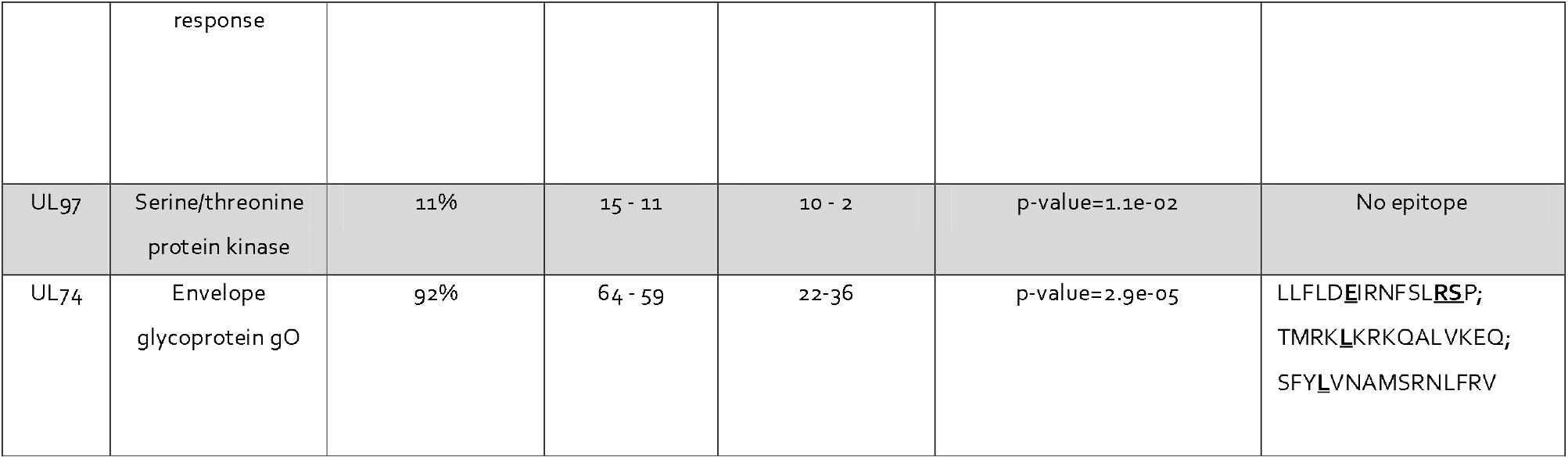
The table shows biological features of the ten HCMV genes under investigation in patients who died. The table shows the % of NS variants mapping to HCMV variable sites, the number of NS vs S MVs found, p-values indicating whether the NS MVs clustered significantly closer than by chance and known and predicted T cell epitopes from the IEDB database which MVs mapped to (in bold and underline the position of the MV in the epitope).

### Patients with poor clinical outcomes have lower lymphocyte counts

The finding that MVs are significantly more likely to occur in regions predicted to be immunogenic led us to explore how immunity might relate to low level mutations. Lymphocyte counts were available from a subset of patients with PID/HSCT (Figure 6A). In patients 1 and 2 (who received SCTs) and patient 10 (who received gene therapy) in the good outcome group, lymphocyte counts recovered quickly after treatment (Figure 6A). In contrast, patient 4, 11, 22 and 23, who died, showed no recovery of lymphocyte count after HSCT. Lymphocyte counts were persistently low in both groups just after HSCT or gene therapy and started to increase at day 100 after transplant. Linear mixed effect modelling showed a significant difference in the counts over time (Figure 6B, p-value < .001) with significant differences in the final lymphocyte counts (good outcome median lymphocyte count: 8.34, 95%CI: 6.69-8.34; poor outcome median lymphocyte count: 0.275, 95%CI: 0.14-1.10).

**Figure 6A:**
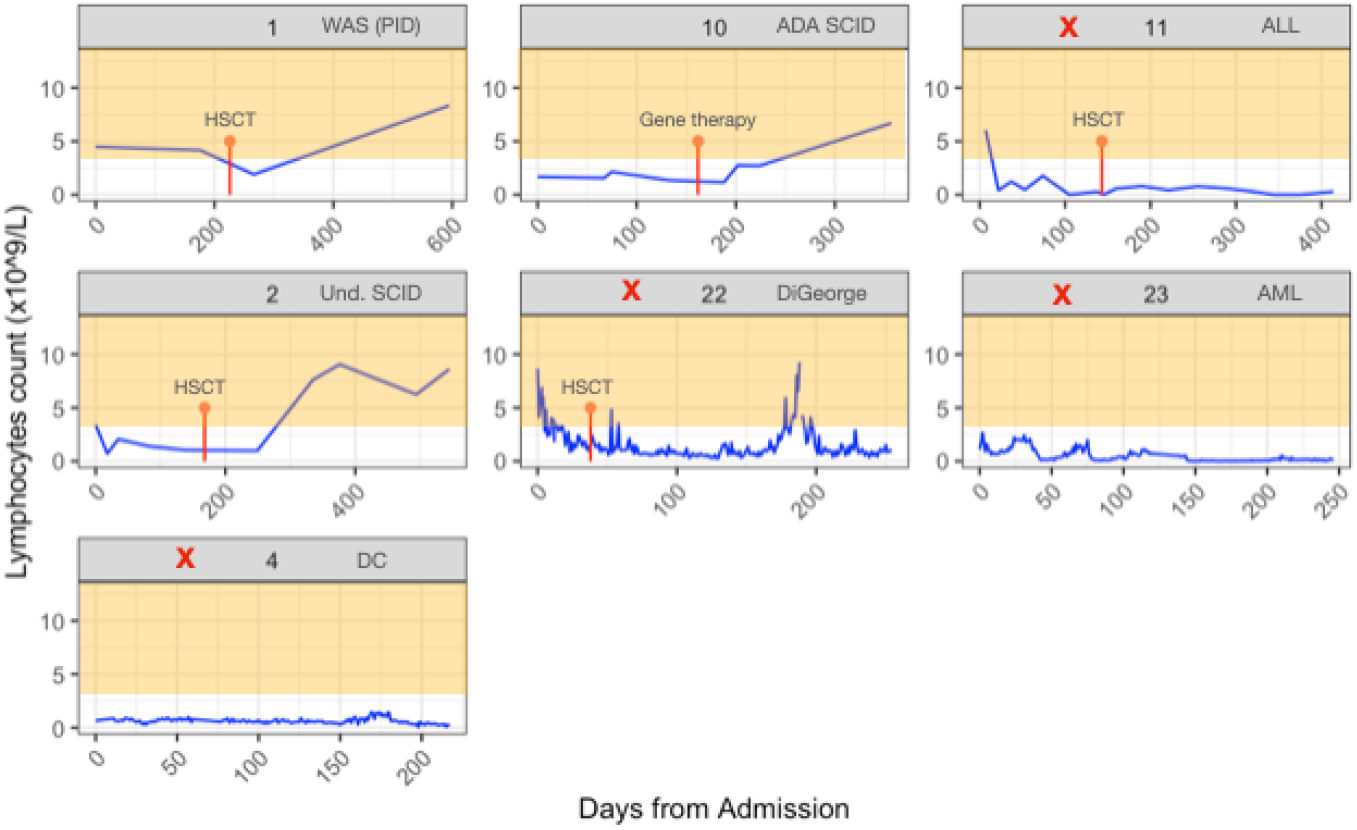
Lymphocyte count (per microliter of blood) overtime in a subset of GOSH patients. Time of HSCT or gene therapy is shown in red. In orange we indicate the healthy lymphocyte count range for children (3-13). Patients with poor outcomes are indicated with a red cross.

**Figure 6B:**
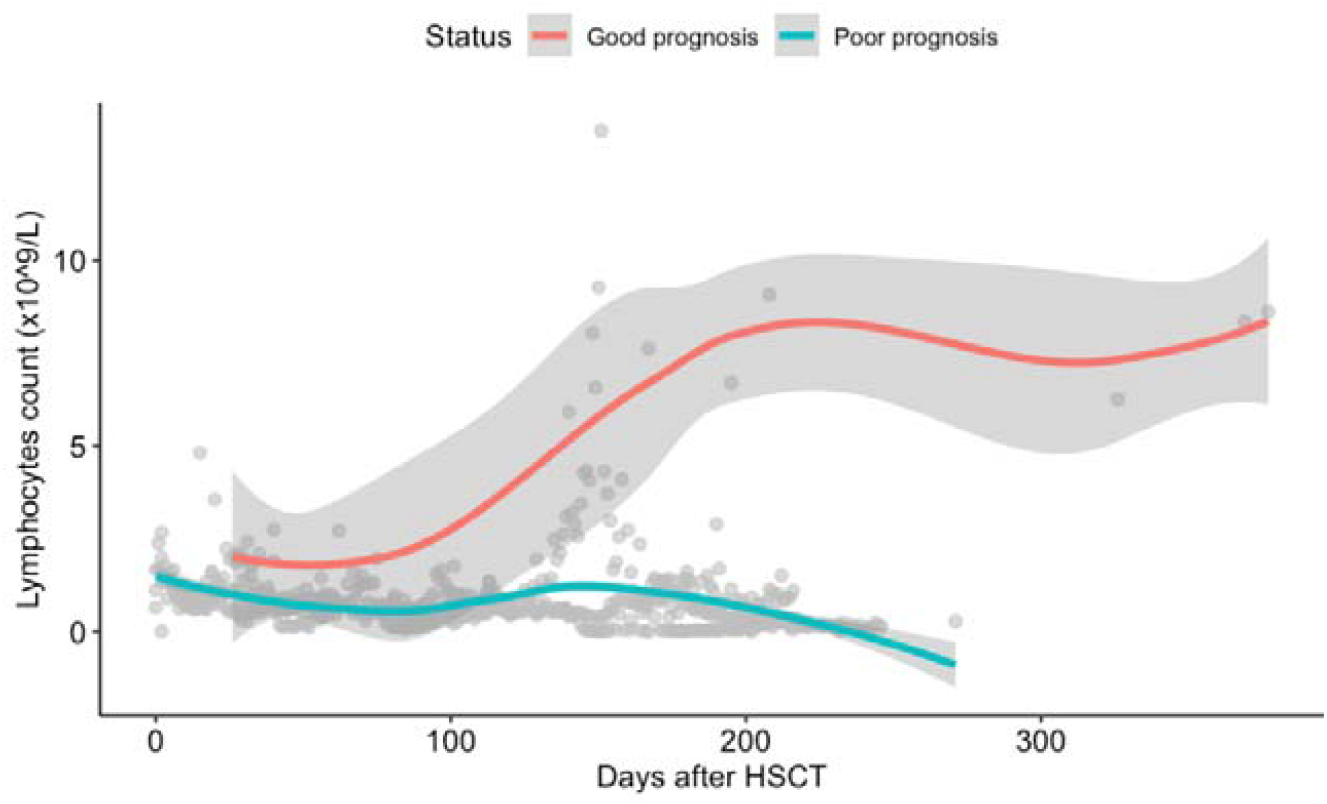
Trend lines (smoothed local regression line using loess) for lymphocyte count for good and poor outcome groups. The grey area represents 95% CI.

Analysing the SOT adult cohort separately, as lymphocyte counts change with age, patient R01-00014 who died also showed persistently lower lymphocyte counts for months after receiving liver transplant as compared with the rest of the SOT cohort who survived (Figure 7A and 7B) (last time point before death for R01-00014 was 0.22, the median in the rest of the SOT patients was 1.61, 95% 0.68-1.61).

**Figure 7A:**
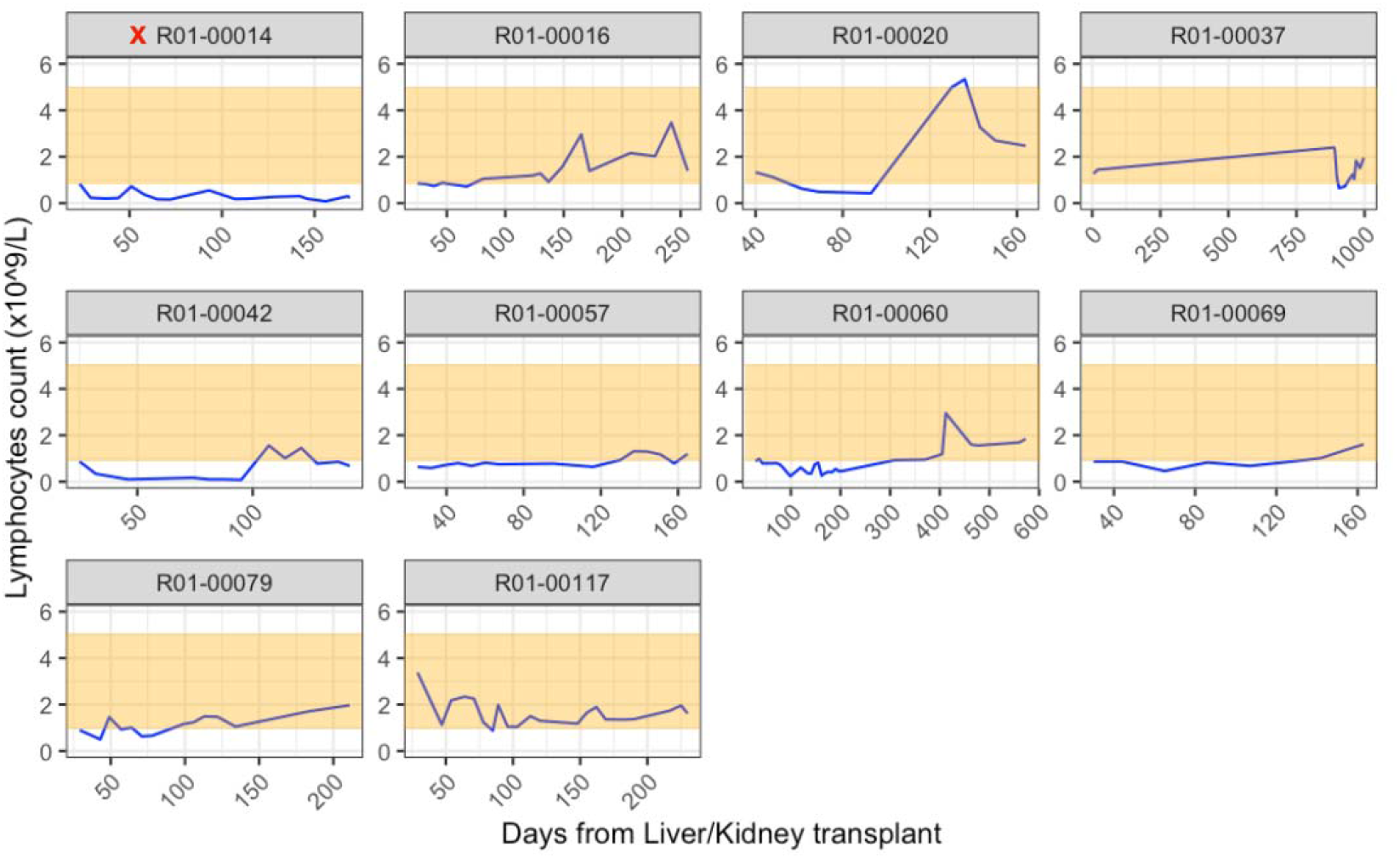
Lymphocyte count (per microliter of blood) overtime in a subset of liver/kidney adult patients. The first time point is taken shortly after kidney/liver transplant. In orange we indicated the healthy lymphocyte count range for adults (0.8-5).

**Figure 7B:**
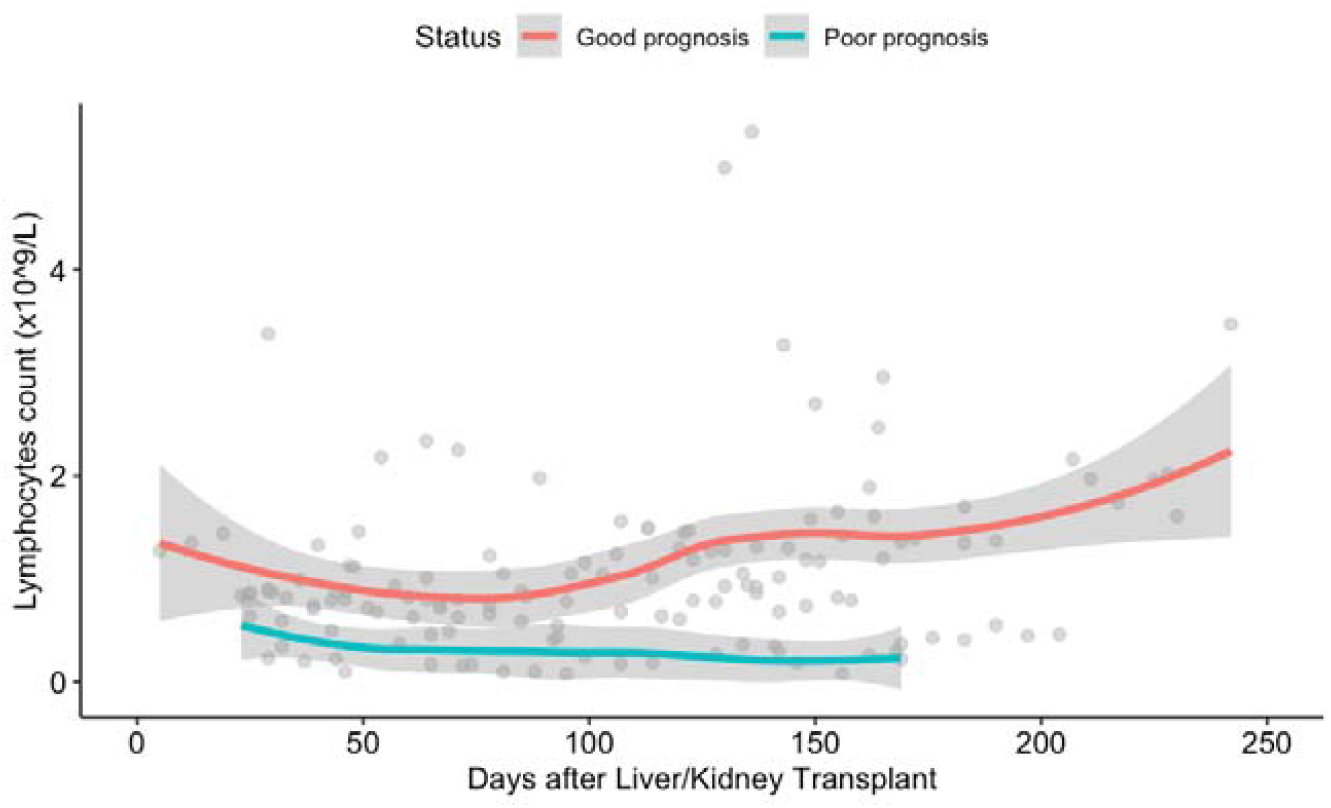
: Trend lines (smoothed local regression line using loess) for lymphocyte count for good and poor outcome groups in a subset of liver/kidney adult patients. The grey area represents 95% CI. X-axis is restricted to 250 days after transplant.

## DISCUSSION

Cytomegalovirus (HCMV) is the most common cause of infection following bone marrow and solid organ transplants (7,20). Active disease significantly increases morbidity and mortality and decreases graft survival in immunosuppressed patients. Pre-emptive therapy and prophylaxis have reduced mortality, but HCMV related organ diseases remain a concern due to treatment side-effects and the rise of drug resistant HCMV. Persistent viraemia is the outcome of the imbalance in the interaction between the virus and the host: HCMV disease occurs when viral replication is uncontrolled in a setting of impaired immune response (8). The mechanisms by which HCMV infection influences transplant outcome are not known (7), but drug resistant HCMV strains and infection with multiple strains have been associated with increased morbidity and mortality (11–14).

To investigate viral factors influencing transplant outcome and mortality, we deep sequenced a total of 150 viruses from 16 children following SCT or SOT and from 35 adult SOT recipients. Nine patients died with persistent HCMV viraemia, whilst the remaining 42 cleared their HCMV. Multiple-strain infections are common in immune-compromised individuals and in our cohorts. We identified a slightly higher percentage of mixed infections in patients who died (33% vs 26%), but the difference was not significant in our study.

The use of antiviral drugs in the treatment of HCMV-disease perturbs the viral population, selecting for variants in genes that cause drug-resistance. About 25% of the patients analysed in this study showed resistance mutations at various levels in the DNA polymerase UL54 and the protein kinase UL97, which are the major drug targets. Fixed mutations were present in both patients with poor and good outcomes, albeit at lower levels in the latter. In contrast low level variants were almost exclusively present in samples from patients who died. Interestingly, low level resistant variants detected in two patients with good outcomes, quickly rose to fixation, whereas those detected in three patients with poor outcomes for whom longitudinal samples were available persisted at low level in the subsequent samples. Thus, the finding of low-level resistance mutations should trigger repeat testing to better define the phenotype as well as to identify early resistance mutations that may become fixed and require treatment change. Compared to traditional PCR and Sanger sequencing, NGS can detect low-level resistance mutations at high resolution, enabling the detection of evolving virus populations in immunocompromised individuals selected under anti-viral treatment (11,21–23).

These data and previous observations confirm that HCMV is highly stable at consensus level in immunocompromised patients with very few substitutions observed over time in single-strain infections (0-25 substitutions) (12,23,24). To further investigate the apparently greater low-level mutability in some patients, we used a machine learning approach to attempt to discriminate between patients with good and poor clinical outcomes. Comparing single-strain infected patients who died with those who survived we identified the presence of low level (<50%) variants in one or more of 10 genes, including UL54 and UL97 as discriminatory between the two groups. Notwithstanding the opportunistic nature of the samples available, we were able to detect this signature on average 84 days before death and <100 days post-transplant and in all cases the signature was present in the first available sample from patients.

Most of the HCMV genome is under purifying selection (18), however 5/10 genes from the signature identified had a higher proportion of NS variants than expected from pan-genome analysis and mapped to loci known to be variable in HCMV. Variable loci clustered more than expected by chance, hinting at the possibility of local positive selection a hallmark of immune epitopes (Table 5) and indeed, in seven out of 10 of the signature protein genes MVs mapped to known HCMV T cell epitopes. There might be several reasons why these variants remain at low frequency. Although variation at consensus sequence level is rare due to the proofreading activity of the viral DNA polymerase (25), one possibility is that low level variation in these epitopes occurs normally, but is cleared by functional T cell immunity. Variants are unlikely to confer increased fitness, rising to fixation only in circumstances where they enable evasion of prevailing immunity. In the absence of functional T cell immunity, as in the cases with poor outcomes described here, we postulate that variants arising in epitopes are able to persist at low level long enough to allow detection by deep sequencing. Mutations arising in loci that confer resistance may rise to fixation in the presence of the drug. However, there is evidence that GCV resistance mutations are not evenly distributed in different cell compartments (26,27) and it is possible that the presence of low level virus subpopulations with antiviral resistance represent virus sequestered in certain cell types.

Taken together the data hint at the possibility that dysregulated immunity in some way contributes to the accumulation of low-level variants. There is evidence from early studies that recovery of CD8+ T cells and CD4+ T cells is a positive predictor for prevention of mortality in HCMV-disease (28–30). Restoration of HCMV-specific CTL response (class I MHC-restricted specific CD8+ CTL) may require an extended time after transplant in some patients, and such patients are at increased risk of developing severe HCMV disease. In our study, we were not able to obtain measurement of T cell function, largely because the peripheral blood lymphocyte subset counts were too low for the assays used. Instead, we analysed lymphocyte counts as proxy for lymphocyte function in a subset of 7 children for whom data was available. None of the four patients (three post SCT and one with PID) who died had measurable lymphocyte counts and all harboured viral variants in the ten genes as described above. In contrast three subjects with a good outcome (two post HSCT and one after gene therapy) for whom we had data showed good lymphocyte count recovery. Thus, the detection of persistent low-level HCMV mutations following SCT may be a biomarker of poor immune reconstitution and consequent poor outcome of HCMV infection. Although fatal HCMV disease is less common in SOTs, it is interesting that patient R01-00014 who died with disseminated HCMV showed a similar signature to the HSCT patients who died, suggesting that similar processes may underlie fatal HCMV disease irrespective of transplant type. In this opportunistically collected sample set, we did not always have samples early on in HCMV infection. Notwithstanding, the viral signature was present, in all cases in the first sample tested, including at day nine following transplant in one patient. In all cases the signature was detected <100 days after transplant, i.e., before T cell recovery is expected, thus providing a potential early biomarker for failure of engraftment and poor outcome of HCMV infection. Since low level resistance mutations which later rise to fixation can occur in the good prognosis group, repeated testing to demonstrate persistent low level variants is likely to increase specificity. The treatment of patients with HCMV infections that persist in the face of antiviral treatment is challenging. A biomarker that provides an early indication of the likely failure of pharmacological approaches, could provide timely signposting of the likely need for rescue cell-based therapies or even repeat HSCT to achieve control of HCMV in these patients.

This study represents a set of preliminary observations based on a limited number of patients especially in the poor outcome group. The findings now need to be tested prospectively in a larger group of patients. A further limitation is that the biological basis for these observations is not known although we speculate as to a possible explanation. Despite the availability of effective antiviral prophylaxis and treatment, HCMV remains a serious infection, particularly in the context of congenital or acquired persistent poor T cell numbers and function. In this context, the development of antiviral resistance is more common, and the prognosis can be poor. Routine use of next generation sequencing for HCMV resistance in refractory patients could potentially detect significant resistance at earlier timepoints. At the same time, repeated detection of MVs may prove to be a useful biomarker for poor response to drug treatment alone and identify patients, including where there are insufficient cells present for functional T cell assays, for whom non-pharmaceutical rescue therapies may be needed.

## METHODS

### Sample collection and ethics

#### Great Ormond Street Hospital samples

Whole blood samples were stored at Great Ormond Street Hospital for Children (GOSH) at -80C. These residual samples were collected as part of the standard clinical care at GOSH, and subsequently approved for research use through the UCL Partners Infection DNA Bank by the NRES Committee London Fulham (REC reference: 12/LO/1089) and West Midlands Black Country Research Ethics Committee (REC reference: 18/WM/0186). All samples were anonymised. Informed patient consent was not required. Nucleic acid was enriched using custom baits and sequenced as previously described (11,12,31).

#### Royal Free London samples

Samples were collected as part of the Wellcome collaborative grant 204870/Z/16/Z UKRI). UCL17-0008 Analysis of Cytomegalovirus Pathogenesis in Solid Organ Transplant Patients approved by the NRES Committee London Queens Square Ethics Committee (REC reference 17/LO/0916). Nucleic acid was enriched using custom baits and sequenced as previously described (11,12,31).

### Data availability

Data deposition: Raw sequencing data for HCMV have been deposited in the European Nucleotide Archive (project accession no. PRJEB12814 and PRJEB55677 for GOSH patients and PRJEB55701 for SOT WT patients).

### Statistical analysis

#### Bioinformatics analysis

All sequences were analysed by the same methods. Reads were trimmed and QC using Trimgalore (https://www.bioinformatics.babraham.ac.uk/projects/trim_galore/) and then mapped to the Merlin strain (GenBank Id: NC_006273.2) using BBmap (https://jgi.doe.gov/data-and-tools/bbtools/bb-tools-user-guide/bbmap-guide/). Variants were called using Varscan version 2. All non-fixed variants were included in the analysis (frequency cut-off 2%). Diversity calculations have been described elsewhere (12) and code is available here https://github.com/ucl-pathgenomics/NucleotideDiversity. For haplotype reconstruction, we used HaROLD, which uses co-varying variant frequencies in a probabilistic framework. Validation and applications are described here (32) and programs can be found here https://github.com/ucl-pathgenomics/HaROLD.

#### Feature selection

We created a dataset where for each sample we had genes as categorical variables and presence of MVs was indicated as 1/0. Genes with only one mutation in one sample were filtered out. We implemented a gene selection algorithm to evaluate the importance of the presence of low-level variants in a specific gene using Python scikit-learn library (33). Gene selection was done according to the k highest scores (sklearn.feature_selection.SelectKBest with chi-square statistics for classification). Data were split into train/test (70% train, 30% test) 1000 times and, to avoid bias due to longitudinal sampling, we used a Leave-One-Out Cross-Validation (LOOCV) procedure, in particular the shuffle-group-out cross-validation iterator implemented in scikit-learn library (sklearn.model_selection.GroupShuffleSplit) which provides randomized train/test indices to split data according to patient variable. Genes were selected based on 1) top 10 with the highest k-score 2) adjusted -log(p-value) of 1 and k-score of 8.

#### Regression model accuracy and probability

A generalised logistic model (R function glm, family binomial) for implemented to test the accuracy of the 10-genes model in predicting the clinical outcome. The ‘predict()’ function was used to predict the response value of each observations as probability to be part of the poor prognosis group. ROC curves (function ‘roc()’) were used to show the sensitivity/specificity for the binary classifier and the area under the curve (AUC) was also calculated.

The results were compared with a model using only the two resistance genes, UL54 and UL97 using an ANOVA test (likelihood-ration, LR).

#### Biology of the genes

Genes were annotated and tested for drug resistance using the R package cmvdrg (16). T cell epitopes for HCMV were extracted from the Immune Epitope Database and Analysis Resource (IEDB). Lymphocytes counts were compared with a mixed effect regression model in R.

## Data Availability

Raw sequencing data have been deposited in the European Nucleotide Archive (project accession no. PRJEB12814 and PRJEB55677 for GOSH patients and PRJEB55701 for SOT WT patients).

## Abbreviations

HCMV: Human Cytomegalovirus
Kbp: kilobase pairs
NGS: Next Generation Sequencing
WGS: Whole Genome Sequencing
SOT: Solid organ transplant
HSCT: haematopoietic stem cell transplantation
PID: primary immunodeficiency
BMT: bone marrow transplant
GCV: ganciclovir
FOS: foscarnet
CDV: cidofovir
MV: minority variant

## ACKNOWLEDGMENTS

We are grateful for the excellent help from the Pathogen Genomics stream of UCL Genomics.

## SUPPLEMENTARY FIGURES

**Figure 1 – figure supplement 1:**
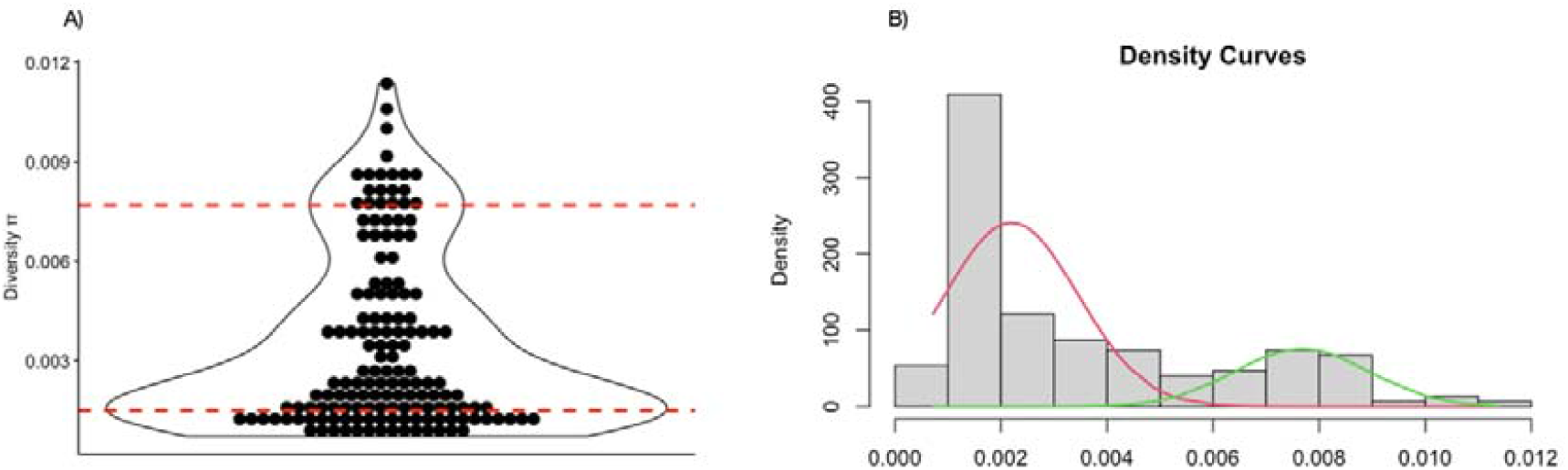
: Distribution of diversity values for all samples (A). Red dashed lines represent the two modes of the bi-modal distribution. The estimated modes were used to create a mixture of Gaussian distributions as shown in the plot (B).

**Table 2 – figure supplement 1:**
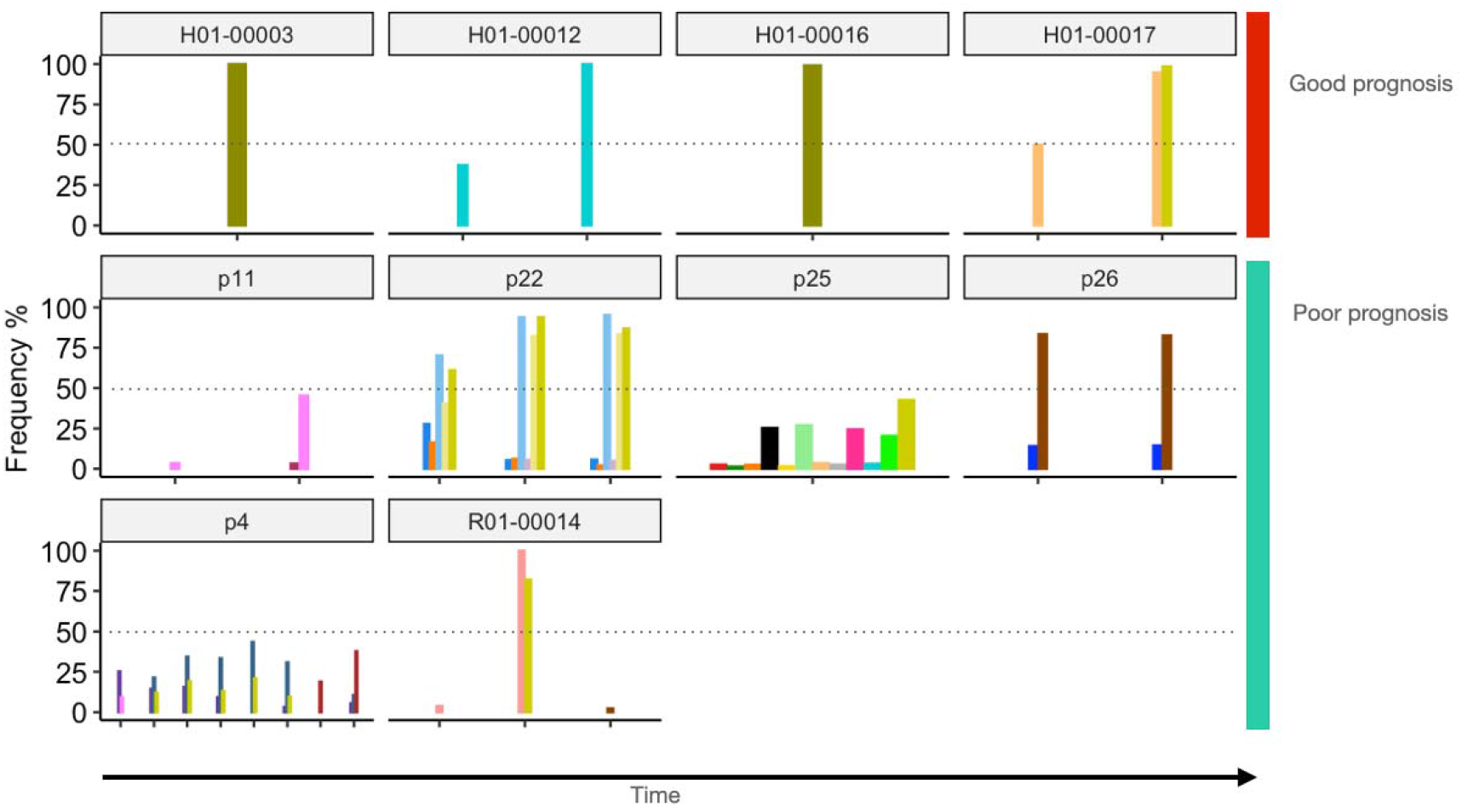
Resistance variants overtime (x-axis) in patients with good (red) and poor (blue) clinical outcome. Variants are considered low-level if the frequency (y-axis) was below 50%.

**Figure 4 – supplement figure 1:**
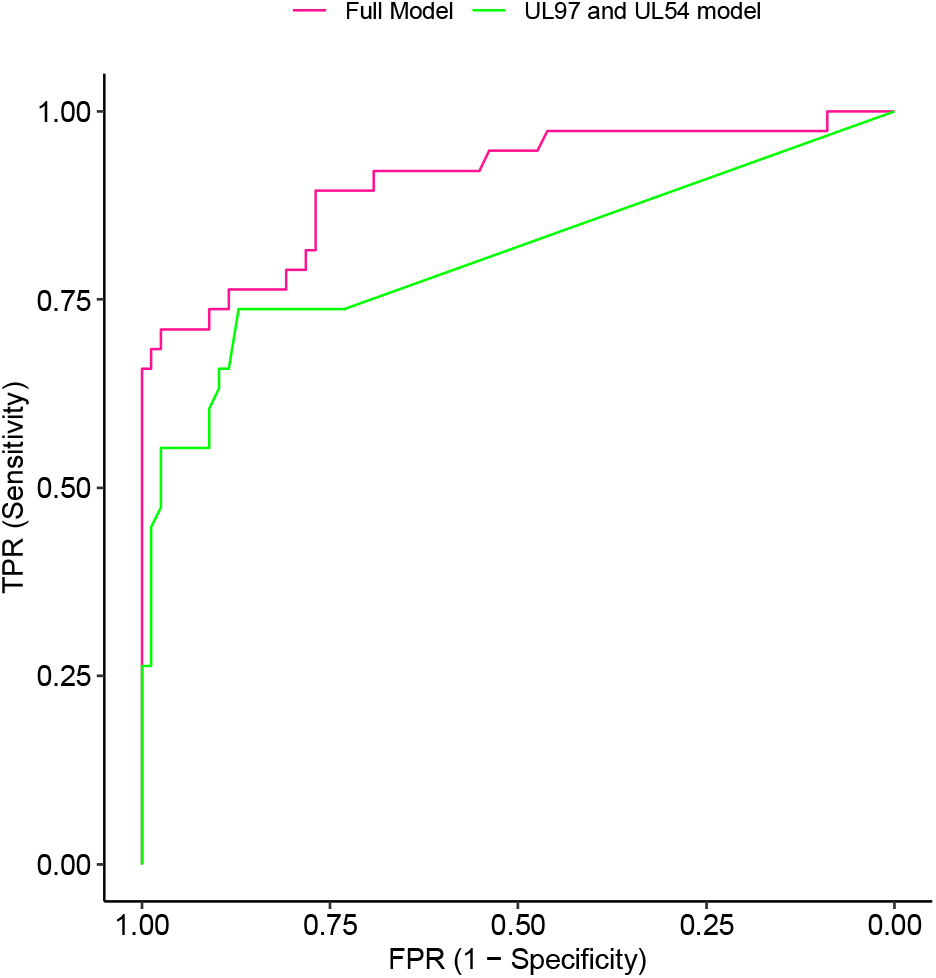
ROC curves with confidence intervals (95%) for two predictive models discriminating between samples from patients who died and survivors including all samples from single and mixed infections. AUC for the full model (including MVs in the 10 candidate genes) was 0.91 (red ROC curve). AUC for the drug resistance genes model (including genes UL54 and UL97) was 0.81 (green ROC curve). The two models were significantly different (p-value < 0.001, Anova).

## REFERENCES

1. Cannon MJ, Schmid DS, Hyde TB. Review of cytomegalovirus seroprevalence and demographic characteristics associated with infection. Reviews in Medical Virology. 2010;20(4):202–13.

2. Zuhair M, Smit GSA, Wallis G, Jabbar F, Smith C, Devleesschauwer B, et al. Estimation of the worldwide seroprevalence of cytomegalovirus: A systematic review and meta-analysis. Reviews in Medical Virology. 2019;29(3):e2034.

3. Green ML, Leisenring W, Xie H, Mast TC, Cui Y, Sandmaier BM, et al. Cytomegalovirus viral load and mortality after haemopoietic stem cell transplantation in the era of pre-emptive therapy: a retrospective cohort study. The Lancet Haematology. 2016 Mar 1;3(3):e119–27.

4. Ramanan P, Razonable RR. Cytomegalovirus Infections in Solid Organ Transplantation: A Review. Infect Chemother. 2013 Sep 27;45(3):260–71.

5. Kotton CN, Kumar D, Caliendo AM, Huprikar S, Chou S, Danziger-Isakov L, et al. The Third International Consensus Guidelines on the Management of Cytomegalovirus in Solid-organ Transplantation. Transplantation. 2018 Jun;102(6):900–31.

6. Boeckh M, Ljungman P. How we treat cytomegalovirus in hematopoietic cell transplant recipients. Blood. 2009 Jun 4;113(23):5711–9.

7. Kotton CN. Management of cytomegalovirus infection in solid organ transplantation. Nat Rev Nephrol. 2010 Dec;6(12):711–21.

8. Bruminhent J, Razonable RR. Advances in drug therapies for cytomegalovirus in transplantation: a focus on maribavir and letermovir. Expert Opinion on Orphan Drugs. 2020 Oct 2;8(10):393–401.

9. Shmueli E, Or R, Shapira MY, Resnick IB, Caplan O, Bdolah-Abram T, et al. High Rate of Cytomegalovirus Drug Resistance Among Patients Receiving Preemptive Antiviral Treatment After Haploidentical Stem Cell Transplantation. The Journal of Infectious Diseases. 2014 Feb 15;209(4):557–61.

10. van der Beek MT, Marijt EW, Vossen AC, van der Blij-de Brouwer CS, Wolterbeek R, Halkes CJ, et al. Failure of Pre-Emptive Treatment of Cytomegalovirus Infections and Antiviral Resistance in Stem Cell Transplant Recipients. Antiviral Therapy. 2012 Jan 1;17(1):45–51.

11. Houldcroft CJ, Bryant JM, Depledge DP, Margetts BK, Simmonds J, Nicolaou S, et al. Detection of Low Frequency Multi-Drug Resistance and Novel Putative Maribavir Resistance in Immunocompromised Pediatric Patients with Cytomegalovirus. Front Microbiol [Internet]. 2016 [cited 2020 May 27];7. Available from: https://www.frontiersin.org/articles/10.3389/fmicb.2016.01317/full

12. Cudini J, Roy S, Houldcroft CJ, Bryant JM, Depledge DP, Tutill H, et al. Human cytomegalovirus haplotype reconstruction reveals high diversity due to superinfection and evidence of within-host recombination. PNAS. 2019 Mar 19;116(12):5693–8.

13. Coaquette A, Bourgeois A, Dirand C, Varin A, Chen W, Herbein G. Mixed Cytomegalovirus Glycoprotein B Genotypes in Immunocompromised Patients. Clinical Infectious Diseases. 2004 Jul 15;39(2):155–61.

14. Lisboa LF, Tong Y, Kumar D, Pang XL, Åsberg A, Hartmann A, et al. Analysis and clinical correlation of genetic variation in cytomegalovirus. Transplant Infectious Disease. 2012;14(2):132–40.

15. Chemaly RF, Chou S, Einsele H, Griffiths P, Avery R, Razonable RR, et al. Definitions of Resistant and Refractory Cytomegalovirus Infection and Disease in Transplant Recipients for Use in Clinical Trials. Clinical Infectious Diseases. 2019 Apr 8;68(8):1420–6.

16. Charles OJ, Venturini C, Breuer J. cmvdrg - An R package for Human Cytomegalovirus antiviral Drug Resistance Genotyping [Internet]. bioRxiv; 2020 [cited 2022 Apr 5]. p. 2020.05.15.097907. Available from: https://www.biorxiv.org/content/10.1101/2020.05.15.097907v1

17. Pang J, Slyker JA, Roy S, Bryant J, Atkinson C, Cudini J, et al. Mixed cytomegalovirus genotypes in HIV-positive mothers show compartmentalization and distinct patterns of transmission to infants. Stanley M, Akhmanova A, Ramchandar N, editors. eLife. 2020 Dec 31;9:e63199.

18. Lassalle F, Depledge DP, Reeves MB, Brown AC, Christiansen MT, Tutill HJ, et al. Islands of linkage in an ocean of pervasive recombination reveals two-speed evolution of human cytomegalovirus genomes. Virus Evolution. 2016 Jan 1;2(1):vew017.

19. Suárez NM, Musonda KG, Escriva E, Njenga M, Agbueze A, Camiolo S, et al. Multiple-Strain Infections of Human Cytomegalovirus With High Genomic Diversity Are Common in Breast Milk From Human Immunodeficiency Virus–Infected Women in Zambia. The Journal of Infectious Diseases. 2019 Jul 31;220(5):792–801.

20. Hiwarkar P, Gaspar HB, Gilmour K, Jagani M, Chiesa R, Bennett-Rees N, et al. Impact of viral reactivations in the era of pre-emptive antiviral drug therapy following allogeneic haematopoietic SCT in paediatric recipients. Bone Marrow Transplant. 2013 Jun;48(6):803–8.

21. Chou S, Ercolani RJ, Sahoo MK, Lefterova MI, Strasfeld LM, Pinsky BA. Improved Detection of Emerging Drug-Resistant Mutant Cytomegalovirus Subpopulations by Deep Sequencing. Antimicrob Agents Chemother. 2014 Aug;58(8):4697–702.

22. Guermouche H, Burrel S, Mercier-Darty M, Kofman T, Rogier O, Pawlotsky JM, et al. Characterization of the dynamics of human cytomegalovirus resistance to antiviral drugs by ultra-deep sequencing. Antiviral Research. 2020 Jan 1;173:104647.

23. Suárez NM, Blyth E, Li K, Ganzenmueller T, Camiolo S, Avdic S, et al. Whole-Genome Approach to Assessing Human Cytomegalovirus Dynamics in Transplant Patients Undergoing Antiviral Therapy. Front Cell Infect Microbiol [Internet]. 2020 [cited 2020 Aug 18];10. Available from: https://www.frontiersin.org/articles/10.3389/fcimb.2020.00267/full#h3

24. Hage E, Wilkie GS, Linnenweber-Held S, Dhingra A, Suárez NM, Schmidt JJ, et al. Characterization of Human Cytomegalovirus Genome Diversity in Immunocompromised Hosts by Whole-Genome Sequencing Directly From Clinical Specimens. The Journal of Infectious Diseases. 2017 Jun 1;215(11):1673–83.

25. Renzette N, Gibson L, Jensen JD, Kowalik TF. Human cytomegalovirus intrahost evolution—a new avenue for understanding and controlling herpesvirus infections. Current Opinion in Virology. 2014 Oct 1;8:109–15.

26. Eckle T, Prix L, Jahn G, Klingebiel T, Handgretinger R, Selle B, et al. Drug-resistant human cytomegalovirus infection in children after allogeneic stem cell transplantation may have different clinical outcomes. Blood. 2000 Nov 1;96(9):3286–9.

27. Frange P, Boutolleau D, Leruez-Ville M, Touzot F, Cros G, Heritier S, et al. Temporal and spatial compartmentalization of drug-resistant cytomegalovirus (CMV) in a child with CMV meningoencephalitis: Implications for sampling in molecular diagnosis. Journal of Clinical Microbiology. 2013;51(12):4266–9.

28. Griffiths P, Reeves M. Pathogenesis of human cytomegalovirus in the immunocompromised host. Nat Rev Microbiol. 2021 Dec;19(12):759–73.

29. Reusser P, Riddell S, Meyers J, Greenberg P. Cytotoxic T-lymphocyte response to cytomegalovirus after human allogeneic bone marrow transplantation: pattern of recovery and correlation with cytomegalovirus infection and disease. Blood. 1991 Sep 1;78(5):1373–80.

30. Quinnan GV, Kirmani N, Rook AH, Manischewitz JF, Jackson L, Moreschi G, et al. Cytotoxic T Cells in Cytomegalovirus Infection. New England Journal of Medicine. 1982 Jul 1;307(1):7–13.

31. Depledge DP, Palser AL, Watson SJ, Lai IYC, Gray ER, Grant P, et al. Specific Capture and Whole-Genome Sequencing of Viruses from Clinical Samples. PLOS ONE. 2011 Nov 18;6(11):e27805.

32. Pang J, Venturini C, Tamuri AU, Roy S, Breuer J, Goldstein RA. Haplotype assignment of longitudinal viral deep-sequencing data using co-variation of variant frequencies. bioRxiv. 2020 Aug 27;444877.

33. Pedregosa F, Varoquaux G, Gramfort A, Michel V, Thirion B, Grisel O, et al. Scikit-learn: Machine Learning in Python. Journal of Machine Learning Research. 2011;12(85):2825–30.

