## Supplementary material for "Persistent low-level variants in a subset of HCMV genes are highly predictive of poor outcome in immunocompromised patients with cytomegalovirus infection": Table 1

| **Patient** | **N of samples (after QC)** | **Underlying diagnosis** | **Peak HCMV (genome copies/ml )** | **Duration of viraemia (days at > 10^4^ gc/ml)** | **Anti-viral treatment** | **Clinical outcomes** |
| --- | --- | --- | --- | --- | --- | --- |
| **1** | 1 | Wiskott-Aldrich syndrome | 505906 | 36 | ACV, GCV, VGC | Well after gene therapy |
| **2** | 4 | Undefined severe combined immunodeficiency | 99069 | 26 | ACV, FOS, GCV | Well after SCT. Has nephrotic syndrome unrelated to SCID |
| **3** | 1 | B-acute lymphoblastic leukemia | 2134470 | 28 | FOS, GCV | Well after SCT |
| **4** | 9 | Dyskeratosis congenita | 65611500 | 200 | FOS, GCV, CDV, MBV, LEF, ART, CMV-IVIG | Did not have SCT. Died of CMV colitis and CMV pneumonities |
| **10** | 6 | ADA SCID | 3965090 | 45 | ACV, FOS, GCV, CDV, palivizumab | Well after gene therapy |
| **11** | 7 | Acute Lymphoblastic leukaemia | 1257520 | 274 | GCV, FOS, VGC, ACV | Marrow transplant, died due to gram negative sepsis after CART cell therapy |
| **16** | 8 | RAG2 SCID | 1016020 | 37 | GCV, FOS | Presented with CMV pneumonitis and died after SCT of CMV pneumonitis |
| **17** | 5 | Acute Lymphoblastic leukaemia | 250965 | 62 | GCV, CDV | PBSCT transplant died after thrombotic microangiopathy after SCT |
| **18** | 4 | Undefined immunodeficiency | 2014670 | 90 | GCV, VGC | Noted to have absent B and NK cells, CD3 lymphopenic and low IgG despite SCIG therapy. Multiple chest issues, including CMV pneumonitis and ongoing persistent CMV viraemia. |
| **19** | 1 | Undefined immunodeficiency | 3795840 | 81 | ACV, FOS, GCV, VGC | Chronic lung diseases. EBV driven lymphoproliferative disease. Well after SCT |
| **20** | 2 | Chronic granulomatous disease (p40 deficiency) | 52855 | 23 | VGC | LRD renal transplant (father) Nov 2015. CMV viraemia, acute T-cell mediated rejection. Received methylpred, plasma exchange, rituximab, IVIG. |
| **22** | 3 | DiGeorge syndrome | 16721700 | 157 | FOS, GCV, CDV, palivizumab | Died after thymic transplant due to chronic lung disease. Persistently high levels CMV viraemia |
| **23** | 3 | Acute myeloid leukaemia | 11728700 | 57 | ACV, FOS | Died after SCT due to disease relapse |
| **24** | 2 | Heart transplant | 393192 | 12 (but antivirals >21 days) | GCV, V-GCV | On-going treatment |
| **25** | 1 | Acute myeloid leukaemia | 9160000 | 90 | GCV, FOS, VGC, ACV | Death |
| **26** | 2 | DiGeorge syndrome | >20000000 | 27 | CDV, FOS, GCV, | Too unwell to receive thymus transplant due to disseminated CMV disease. Transferred back home for palliation |

**Table 1. Patients characteristics (paediatric patients from Great Ormond Street Hospital).**
